# Breast Cancer Detection with Topological Deep Learning

**DOI:** 10.1101/2025.10.15.25338098

**Authors:** Brighton Nuwagira, Adrian Rodriguez, Qiwei Li, Baris Coskunuzer

## Abstract

In this paper, we investigate the integration of topological data analysis (TDA) techniques with deep learning (DL) models to improve breast cancer diagnosis from ultrasound images. By leveraging persistent homology, a TDA method that captures global structural patterns, we enrich the local spatial features typically learned by DL models. We incorporate topological features into various pre-trained architectures, including CNNs and vision transformers (VTs), aiming to enhance screening accuracy for breast cancer, which remains the most common cancer among women.

Experiments on publicly available ultrasound datasets demonstrate that combining CNNs and VTs with topological features consistently yields statistically significant performance improvements. Notably, this approach also helps to address challenges faced by DL models, such as interpretability and reliance on large labeled datasets. Furthermore, we generalize the Alexander duality theorem to cubical persistence, showing that persistent homology remains invariant under sublevel and superlevel filtrations for image data. This advancement reduces computational costs, making TDA methods more practical for image analysis.

## 1. Introduction

Medical image analysis plays a pivotal role in modern healthcare, revolutionizing diagnostics, treatment planning, and patient care. The application of deep learning (DL) models in medical image analysis has drastically transformed diagnostic procedures, significantly improving disease detection and categorization [44, 60]. DL models excel in extracting intricate patterns from complex medical images like Magnetic resonance imaging (MRI) scans, computed tomography (CT) scans, and X-rays, empowering healthcare professionals with enhanced capabilities. They have demonstrated exceptional performance in tasks such as tumor detection, organ segmentation, and anomaly identification, reducing human error and expediting analyses [79]. However, challenges such as data scarcity, interpretability, and clinical validation remain areas of active research and development [6, 64].

On the other hand, topological machine learning (ML) presents promising solutions to address the limitations of DL models. Its primary strength lies in its ability to systematically analyze complex spatial relationships within images, surpassing traditional pixel-based analysis [65, 66]. Topological methods delve into the inherent structure of data, providing deeper insights into intricate anatomical features and their connections. Unlike DL models, which rely on predefined architectures, topological ML adapts more flexibly to diverse data complexities and irregularities [35]. Moreover, these techniques offer robustness against noise and yield more interpretable outcomes, crucial in medical contexts where transparency and reliability are critical. Furthermore, DL models are adept at capturing intricate local spatial relationships within images, whereas topological data analysis (TDA) methods offer a holistic viewpoint by globally analyzing data, thus providing complementary insights to DL features. By integrating topological insights with DL models, medical image analysis stands to gain improved accuracy, sensitivity to subtle changes, and a comprehensive understanding of biological structures, thus advancing precise diagnostics and treatments.

In this paper, we aim to demonstrate the effectiveness of topological methods in medical image analysis and highlight their synergistic potential with DL models by demonstrating their practical applications. While the incorporation of topological features with graph neural networks (GNNs) has been a very active area of research in graph representation learning [39, 77], the potential synergy of combining these features with DL models in computer vision and medical image analysis remains largely unexplored. With the adaptation of topological features, we aim to enhance the capabilities of DL models for computer-aided diagnosis.

We study the performance of integration of topological features to pre-trained DL models in breast cancer screening, which is the most prevalent cancer in women [70]. Our experiments showed that topological features consistently give a significant boost to the performance of various convolutional neural networks (CNN) and vision transformer (VT) models. Furthermore, the choice of filtration type is the key question in persistent homology (PH) to produce effective topological features. We establish that for image data, an elegant duality pattern emerges, indicating that sub-level and superlevel filtrations produce virtually identical output for cubical persistence. Our contributions can be summarized as follows:

⋄ We investigate the effectiveness of incorporating topological features to enhance DL models for computer-aided diagnosis, showing that this integration results in *statistically significant improvements* in performance across ultrasound and other medical imaging tasks.

⋄ We further demonstrate the value of topological features in addressing critical challenges in breast cancer diagnostics, such as *limited data* and *interpretability*.

⋄ We prove the *Alexander duality theorem* for cubical persistence, showing that sublevel or superlevel filtrations yield virtually identical output for image data. This result significantly enhances the applicability of TDA in image analysis by effectively halving the computation time.

## 2. Related Work

### 2.1. Deep Learning in Breast Cancer Diagnosis

Breast cancer stands as a formidable challenge for women, lacking a definitive cure to date. However, the emergence of DL techniques has introduced promising avenues in breast cancer screening, diagnosis, and prognosis. These methods have demonstrated remarkable efficacy in identifying breast cancer early on, thereby enhancing the prospects of patient survival [57]. Notably, DL surpasses traditional ML approaches by demanding minimal human intervention while achieving comparable levels of feature extraction [61]. Over the years, the main methods to detect breast cancer have been through various methods like mammography [3], histopathology [59], positron emission tomography (PET) [16], digital breast tomosynthesis (DBT) [8], ultrasound [52], CT [43], and MRI [48]. For a thorough review of these approaches, see excellent surveys [1, 32, 36].

Among these approaches, breast ultrasound stands out as a low-cost, user-friendly, radiation-free, portable technique, particularly adept at differentiating between cystic and solid breast masses. Recent advancements in DL methods for breast cancer diagnosis through ultrasound screening have shown considerable promise for clinical implementation, as evidenced by several studies in recent years [20, 25]. In the past couple of years, the latest DL models in image analysis, namely vision transformers (VTs), have demonstrated remarkable success in the field [7, 26, 53]. Furthermore, several recent reports have indicated that DL-based evaluation of breast ultrasound is comparable to that of a human radiologist [23, 62]. In this paper, we aim to demonstrate the efficacy of topological features in enhancing the performance of DL methods in medical imaging, specifically in breast cancer screening using ultrasound images.

### 2.2. TDA in Medical Image Analysis

PH has been quite effective for pattern recognition in image and shape analysis in the past two decades. In medical image analysis, PH produced power results in cell development [47], tumor detection [19], histopathology [51, 76], neuronal morphology [41], brain functionality analysis [12], functional MRI (fMRI) data [55], and genomic data [54]. For a thorough review of TDA methods in biomedicine and medical imaging, see the excellent surveys [65, 67]. For a collection of TDA applications in several domains, see TDA Applications Library [28, 29].

Recently, there has been a surge of interest in topological deep learning within the ML community, showing great potential to augment existing DL methodologies [49, 80]. The effective utilization of topological features has notably enhanced CNN models, particularly in tasks such as image segmentation [17, 58, 69, 75]. Moreover, there is growing recognition of the pivotal role of topological features in diagnostic tasks across various medical domains [31, 40, 42, 68].

In the case of breast cancer detection, recent works have effectively utilized TDA methods for mammogram and MRI screening [50, 74]. While they incorporate TDA output into DL models to regularize the loss function, our approach is more direct. In our study, we directly integrate topological features into DL architectures to improve the latent embeddings to demonstrate their effectiveness, particularly in breast cancer diagnosis.

## 3. Methodology

In this section, we first present the methodology for extracting topological features from an image using cubical persistence. Next, we elaborate on our ML models, wherein we incorporate these features into DL models. Then, we prove Alexander duality result between sublevel-superlevel filtrations for cubical persistence.

### 3.1. Cubical Persistence

PH in TDA reveals patterns in complex datasets by analyzing shapes at multiple resolutions [18]. While effective for various data formats, we focus on its application in image analysis through *cubical persistence*. PH involves three steps: (1) generating nested topological spaces (*Filtration*), (2) capturing topological changes (*Persistence Diagrams*), and (3) transforming these diagrams into ML-ready vectors (*Vectorization*) [24].

#### Step 1 - Constructing Filtrations

Since PH essentially functions as a mechanism for monitoring the progression of topological characteristics within a sequence of simplicial complexes, constructing this sequence stands out as a crucial step. In image analysis, the common approach involves generating a nested sequence of binary images, also known as *cubical complexes*. To achieve this from a given grayscale image χ (with dimensions *r ×s*), grayscale values *γ*_*ij*_ of individual pixels Δ_*ij*_ ⊂ χ are utilized. Specifically, for a sequence of grayscale values (0 = *t*_1_ *< t*_2_ *<* · · · *< t*_*N*_ = 255), a nested succession of binary images χ_1_ ⊂ χ_2_⊂ · · · χ_*N*_ is obtained, where χ_*n*_ ={Δ_*ij*_ ⊂ χ|*γ*_*ij*_ ≤ *t*_*n*_ } (See Figures 1 and 2). In essence, this involves starting with a blank *r* ×*s* image and progressively activating (coloring black) pixels as their color values reach the specified threshold *t*_*n*_. This process, known as *sublevel filtration*, is conducted on χ relative to a designated function (in this instance, grayscale). Alternatively, one can activate pixels in a descending order, referred to as *superlevel filtration*. In other words, let 𝒴_*n*_ = {Δ_*ij*_ ⊂ χ | *γ*_*ij*_ ≥ *s*_*n*_} for (255 = *s*_1_ *> s*_2_ *>* · · · *> s*_*M*_ = 0), and 𝒴_1_ ⊂ 𝒴_2_ ⊂ · · · ⊂ 𝒴_*M*_ is called superlevel filtration.

**Figure 1.**
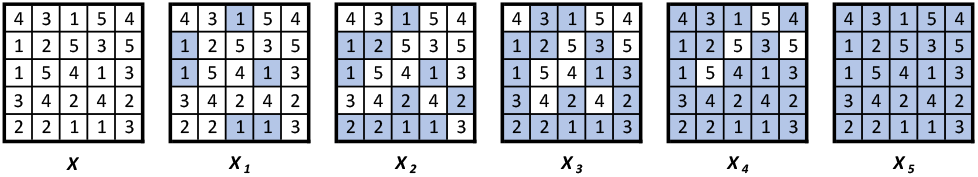
For the 5 *×* 5 image *χ*, **the sublevel filtration** is the sequence of binary images *χ*_1_ ⊂ *χ*_2_ ⊂ *χ*_3_ ⊂ *χ*_4_ ⊂ *χ*_5_.

**Figure 2.**
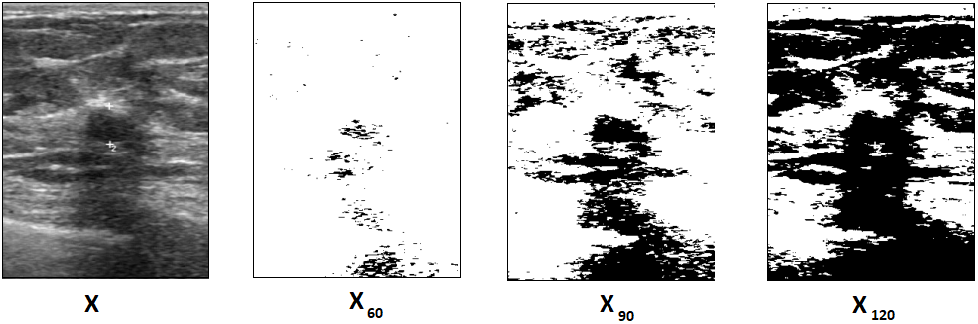
On the left, we have a sample ultrasound image χ from BUS-BRA dataset. Next three images χ_60_, χ_90_, and χ_120_ are binary images in the filtration where in χ_*k*_, pixels with grayscale values ≤ *k* are activated (marked black), while all other pixels remain white.

#### Step 2 - Persistence Diagrams

PH tracks the evolution of topological features within the filtration sequence {χ_*n*_} and represents it as a *persistence diagram* (PD). Specifically, if a topological feature *σ* emerges in χ_*m*_ and vanishes in χ_*n*_ with 1 ≤ *m < n ≤ N*, the thresholds *t*_*m*_ and *t*_*n*_ are termed the *birth time b*_*σ*_ and *death time d*_*σ*_ of *σ*, respectively (*b*_*σ*_ = *t*_*m*_ and *d*_*σ*_ = *t*_*n*_). Hence, PD comprises all such 2-tuples PD_*k*_(χ) = {(*b*_*σ*_, *d*_*σ*_)} where *k* denotes the dimension of the topological features. The duration *d*_*σ*_ *™ b*_*σ*_ is referred to as the *lifespan* of *σ*. Formally, the *k*^*th*^ persistence diagram can be defined as PD_*k*_( χ) = {(*b*_*σ*_, *d*_*σ*_)| *σ* ∈ *H*_*k*_(χ_*n*_)for*b*_*σ*_ *≤ t*_*n*_ *< d*_*σ*_ }, where *H*_*k*_( χ_*n*_) represents the *k*^*th*^ homology group of the cubical complex χ_*n*_. Thus, PD_*k*_(χ) comprises 2-tuples indicating the birth and death times of *k*-dimensional voids {*σ*} (such as connected components, holes, and cavities) in the filtration sequence {χ_*n*_}. For example, for χ in Figure 1, PD_0_(χ) = (1, *∞*), (1, 2), (1, 2), (1, 3) } denotes the connected components, while PD_1_(χ) = {(2, 4), (3, 5), (4, 5)} represents the holes in the corresponding binary images in Figure 1.

#### Step 3 – Vectorizations

Persistence Diagrams (PDs), consisting of collections of 2-tuples, are not inherently practical for utilization with ML tools. Instead, a common strategy is to transform PD information into a vector or function, a process referred to as *vectorization* [5]. A commonly employed function for this purpose is the *Betti function*, which monitors the quantity of *alive* topological features at each threshold. In particular, the Betti function is a step function where *β*_0_(*t*_*n*_) denotes the count of connected components in the binary image χ_*n*_, and *β*_1_(*t*_*n*_) indicates the number of holes (loops) in χ_*n*_. In ML contexts, Betti functions are typically represented as vectors 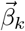 of size *N* with entries *β*_*k*_(*t*_*n*_) for 1 ≤ *n* ≤ *N*, defined as 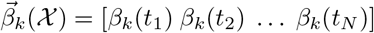. For instance, in Figure 1, we observe 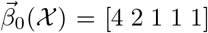, which denotes the count of connected components in the binary images {χ_*i*_}, while 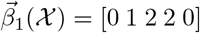, represents the counts of holes in _*i*_, i.e., *β*_0_(1) = 4 signifies the count of components in χ_1_, and *β*_1_(3) = 2 denotes the count of holes (loops) in χ_3_.

There are several approaches for converting PDs into vector representations, such as Persistence Images [2], Persistence Landscapes [11], Silhouettes [14], and kernel-based methods [5]. Here, we use Betti functions due to their computational efficiency and interpretability. Furthermore, Betti functions serve not only as vector representations but also as *sequences* summarizing the number of topological features in the filtration sequence, making them well-suited for use with *transformers* to enhance information extraction. We also employ Persistence Images (Appendix A.5) to study the impact of vectorization on model performance.

### 3.2 ML Models

In this section, we outline our ML models, focusing on approaches that leverage topological features. To evaluate the impact of these features on model performance, we first introduce a baseline model, Topo-ML. This model avoids deep learning, using a traditional ML classifier applied solely to the topological features. Topo-ML is computationally efficient, interpretable, and ideal for scenarios with limited data, thanks to its reliance on direct embeddings. We then present two distinct architectures that integrate these topological features into pre-trained CNN and VT models, enabling deeper exploration of their contributions in more complex frameworks.

#### Topo-ML model

In our basic Topo-ML model, we assessed the standalone performance of topological features using Betti vectors as input. For classification, we employed two standard ML models: eXtreme Gradient Boosting (XG-Boost) [15], Multilayer Perceptron (MLP) [22] and Transformer Encoder [73]. These models are well-suited for handling high-dimensional data, with MLP often excelling in scenarios with large or imbalanced datasets, while XGBoost is known for its robust performance across diverse conditions. A summary of our model structure is shown in Figure 9, and detailed descriptions of our ML procedures, including hyperparameter tuning, are provided in Section 4.

#### Topo-CNN model

In our second model, Topo-CNN (Figure 3), we evaluated the impact of incorporating topological features with pre-trained CNN models. The rationale is that topological features capture the image’s global patterns, while convolutional features focus on localized details. By combining these, we aim to achieve synergistic effects that enhance performance. Specifically, we employed two distinct vectorization methods for topological features to test their effectiveness in deep learning: Betti functions and Persistence Images (PIs). This led to two model variants: Betti-CNN (B-CNN), which uses Betti vectors, and PI-CNN, which incorporates persistence images (Appendix A.5).

**Figure 3.**
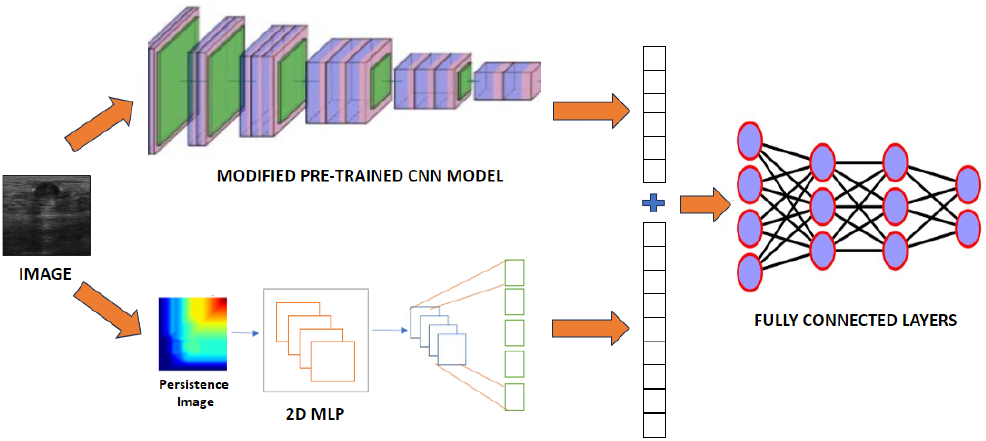
Topo-CNN framework. merges the topological features of images with convolutional vectors derived from a pre-trained CNN model, followed by further processing via a fully connected layer. In our **PI-CNN model**, persistence images are used as topological features and processed through a 2D MLP. In our **Betti-CNN** model, persistence images are replaced with Betti vectors, which are processed through a 1D MLP.

The Topo-CNN model framework involves selecting a CNN backbone from a collection of state-of-the-art architectures (Section 4.2). To analyze the contribution of topological vectors within the DL framework, we employed a simple architecture: convolutional vectors are concatenated with topological vectors after passing through an MLP layer. A flowchart of this model is shown in Figure 3, with hyperparameters and further details provided in Section 4.2.

#### Topo-VT model

In our third model, Topo-VT (Figure 4), we integrate topological features with a vision transformer architecture. This approach leverages the strengths of both global representations, enabling them to complement each other and enhance performance. By facilitating communication between the feature representations of the transformer stages and a transformer-encoded version of topological features, we aim to improve model efficacy.

**Figure 4.**
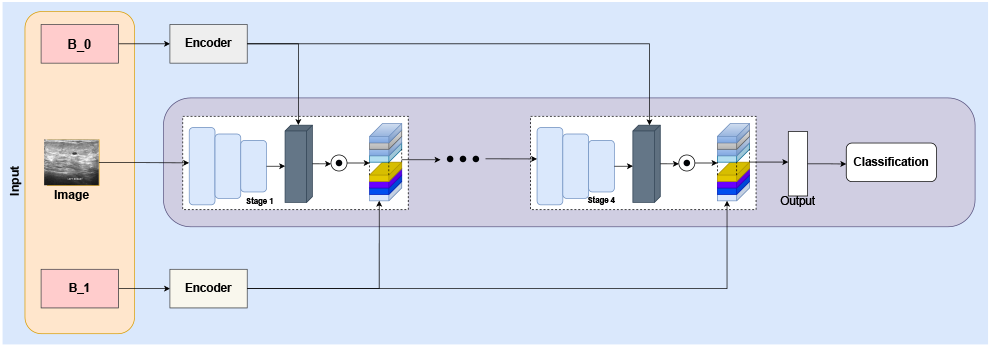
Topo-VT framework. fuses topological features with ViT, applying Betti vector encodings after each stage via cross-attention to incorporate topological insights.

The Topo-VT architecture uses the SwinV2 backbone [45, 46] and introduces Betti vector encodings after each stage to complement the model’s global properties through cross-attention. Each Betti vector 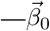 for connected components and 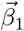 for loops—has its own transformer encoding. We first apply 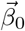 to the feature map after each stage, followed by 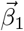 using cross-attention. Given that Betti vectors are sequential data derived from cubical filtration (Section 3.1), transformer encoders are well-suited for integrating topological and transformer-based representations. A flowchart of the model is shown in Figure 4.

### 3.3. Alexander Duality for Cubical Persistence

When employing PH in practical applications, how the filtration is constructed stands out as the most crucial aspect of the process (Section 3.1). For computational efficiency, the sublevel/superlevel filtration method is most preferable, utilizing a suitable filtering function on the data. Particularly in a graph setting, sublevel and superlevel filtrations can yield vastly different outputs depending on the filtering function utilized, and the model’s performance can hinge on the chosen filtration [13]. *Hence, users typically need to examine both filtrations in most TDA applications*.

In the following, we demonstrate that *the sublevel and superlevel filtrations produce virtually identical outputs in complementary dimensions for PH with cubical complexes*. As a result, unlike in the graph setting, the choice between sublevel and superlevel filtration for image analysis has no importance, as long as all relevant dimensions’ outputs are used. We establish this result by adapting *Alexander Duality* to this context. In particular, we prove the following:

#### Theorem 1

*Let* χ *be a cubical complex of dimension d, and f be the filtering function on* χ. *Let* 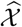 *be its extended image with* 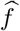 *Then, the persistence diagrams for the sub-level and superlevel filtration of with* 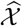 *respect to* 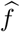*in complementary dimensions are equivalent*.

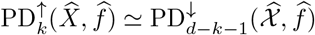

The proof of the theorem and the further details are given in Appendix B. Here, the extended image condition is to make sure that the pixels (or voxels) at the boundary of the image have dark or light color. In particular, if the image has already a dark or light background, this condition is automatically satisfied. To illustrate this duality, we present the Betti curves for an ultrasound image using both sublevel and superlevel filtrations (Figure 5).

**Figure 5.**
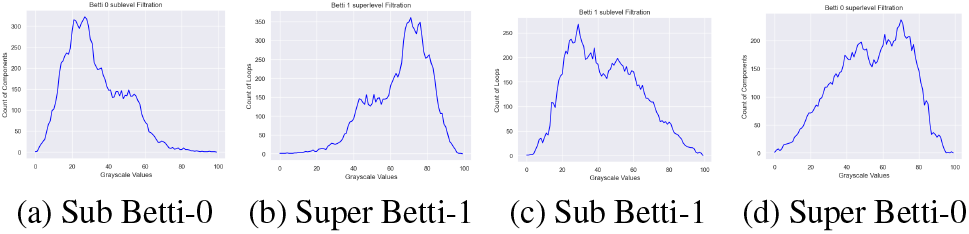
Betti-0 and Betti-1 curves using sublevel and superlevel filtrations for an ultrasound image from the BUS-BRA dataset. As Theorem 1 indicates, sublevel Betti-0 (a) and superlevel Betti-1 (b) curves show mirrored patterns, as do sublevel Betti-1 (c) and superlevel Betti-0 (d) curves.

One way to interpret our result is that if one uses the sublevel filtration for PH with cubical complexes and uses all the relevant dimensions, then *there is no need to check superlevel filtration in the same setting*. In practice, this cuts the computation time by half, and boosts the applicability of TDA methods in image analysis.

## 4. Experiments

### 4.1. Datasets

In our research, we utilized three publicly accessible breast ultrasound datasets (see Table 1). The first dataset, known as BUSI [4], comprises 780 breast ultrasound images collected from 600 women aged 25 to 75 years old. The second ultrasound dataset, BUS-BRA [30], comprises 1875 anonymized images from 1064 female patients. The third ultrasound dataset, MENDELEY [56] consists of 250 breast cancer images, including 100 benign and 150 malignant cases. In addition to these, to show the applicability of our Topo-DL models in different image formats, we employed three other datasets from the MedMNIST collection [78], namely RETINAMNIST (Fundus images), PneuMNIST (Chest X-ray) and BloodMNIST (Blood Cell) (see Table 8).

**Table 1.** Characteristics of breast cancer ultrasound datasets.

| Dataset | Total | Benign | Malignant | Normal | Size |
| --- | --- | --- | --- | --- | --- |
| BUSI | 780 | 437 | 210 | 133 | 500×500 |
| BUS-BRA | 1875 | 1268 | 607 | 0 | 275×375 |
| MENDELEY | 250 | 100 | 150 | 0 | 95×75 |

### 4.2. Experimental Setup

We provide details of our datasets in Table 1. It is important to note that the majority of datasets do not come with a predefined train-test split. Hence, many models utilized their own train-test splits. In this study, we employed 5-fold CV in all datasets except BUSI (3 class) which has a pre-defined split - 10-fold CV. Furthermore, each experiment was run 20 times with different seeds, and the average results were reported in Table 2, and Table 9.

**Table 2.** Topo-CNN results. Performance comparison between vanilla-CNN and Topo-CNN models (Betti-CNN and PI-CNN) for breast ultrasound datasets. For each dataset, the best two accuracy and AUC **improvements** are marked **blue**. In each block, the best performance is **underlined**. The statistical significance of each improvement is given in Tables 13 and 14 in the appendix.

| Backbone | BUSI 2-label |  |  |  |  |  | BUSI 3-label |  |  |  |  |  |
| --- | --- | --- | --- | --- | --- | --- | --- | --- | --- | --- | --- | --- |
|  | Accuracy |  |  | AUC |  |  | Accuracy |  |  | AUC |  |  |
|  | CNN | B-CNN | P-CNN | CNN | B-CNN | P-CNN | CNN | B-CNN | P-CNN | CNN | B-CNN | P-CNN |
| DenseNet121 | 82.53 | 86.22 | 88.05 | 92.04 | 94.18 | 93.65 | 81.29 | 82.96 | 84.24 | **93.97** | **94.87** | **94.07** |
| EfficientNetB0 | 85.01 | 87.57 | 87.42 | 93.34 | 94.83 | 93.02 | **82.69** | **84.08** | **84.10** | 95.01 | 95.17 | 94.04 |
| IncResNetV2 | **73.31** | **86.76** | **87.42** | **81.72** | **94.69** | **94.19** | 80.25 | 82.27 | 83.15 | **93.46** | **94.49** | **93.58** |
| InceptionV3 | 82.61 | 85.00 | 85.62 | 91.08 | 93.16 | 91.98 | 79.77 | 81.91 | 80.69 | 92.91 | 93.91 | 92.23 |
| MobileNetV2 | 83.93 | 86.10 | 87.03 | 93.19 | 93.48 | 92.12 | 82.65 | 83.61 | 80.05 | 94.62 | 94.69 | 91.26 |
| ResNet50 | 85.66 | 87.78 | 88.05 | 93.99 | 94.75 | 93.82 | 83.87 | 84.97 | 85.06 | 95.44 | 95.36 | 94.19 |
| ResNet101 | 87.14 | **89.06** | 88.75 | 95.21 | **95.84** | 94.70 | 84.96 | **85.74** | 85.68 | 95.83 | **95.84** | 95.04 |
| VGG16 | **83.89** | **88.47** | **89.69** | **93.89** | **95.32** | **95.02** | **81.63** | **84.53** | **84.24** | 94.05 | 95.15 | 93.97 |

| Backbone | BUS-BRA |  |  |  |  |  | MENDELEY |  |  |  |  |  |
| --- | --- | --- | --- | --- | --- | --- | --- | --- | --- | --- | --- | --- |
|  | Accuracy |  |  | AUC |  |  | Accuracy |  |  | AUC |  |  |
|  | CNN | B-CNN | P-CNN | CNN | B-CNN | P-CNN | CNN | B-CNN | P-CNN | CNN | B-CNN | P-CNN |
| DenseNet121 | **73.06** | **76.81** | **75.81** | **82.57** | **85.03** | **81.55** | 97.34 | 98.10 | 99.80 | 99.67 | 99.77 | 100.0 |
| EfficientNetB0 | 75.73 | 79.05 | 78.00 | **84.95** | **87.39** | **84.24** | 96.28 | 97.24 | 97.40 | 99.55 | 99.56 | 99.93 |
| IncResNetV2 | 73.77 | 73.31 | 75.03 | 83.54 | 81.72 | 80.17 | **91.90** | **97.54** | **99.40** | **98.62** | **99.72** | **100.0** |
| InceptionV3 | 72.43 | 72.55 | 73.17 | 81.42 | 80.65 | 76.50 | 98.50 | 98.40 | **100.0** | 99.89 | 99.84 | 100.0 |
| MobileNetV2 | 73.67 | 76.05 | 76.52 | 83.27 | 84.65 | 81.74 | 98.42 | 98.14 | 99.60 | 99.86 | 99.78 | 100.0 |
| ResNet50 | 76.47 | 79.38 | 78.96 | 85.32 | 87.23 | 84.25 | 99.66 | 99.12 | 99.60 | 99.98 | 99.96 | 100.0 |
| ResNet101 | 77.15 | 79.32 | **79.96** | 86.01 | 86.96 | 85.13 | 99.62 | 99.22 | 98.80 | 99.98 | 99.97 | 99.93 |
| VGG16 | **69.22** | **75.66** | **74.92** | 80.02 | 83.55 | 78.37 | **97.94** | **99.14** | **99.20** | **99.82** | **99.94** | **100.0** |

We did not use any data augmentation method in this study as the topological features extracted are invariant under rotations or flips and demonstrate robust performance on small and unbalanced datasets. As a result, our TDA models do not require any form of data augmentation or preprocessing. This aspect contributes to the computational efficiency of our models while maintaining a high level of robustness against minor image alterations.

Our implementation is available at the following link: https://github.com/BrightonNuwagira/topo_vt

#### Topological Features

We first resized each ultrasound image χ to 224 × 224, then generated topological feature vectors using a sublevel filtration, χ_1_ ⊂ χ_*N*_, based on grayscale pixel values (see Section 3.1). With *N* = 100 thresholds evenly dividing the color interval [0, 255], we produced 200-dimensional feature vectors by combining Betti vectors 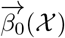 and 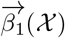, each with 100 dimensions. We also used Persistence Images as vectorization, where the details are given in Appendix A.5.

#### Topo-ML Hyperparameters

In our basic model Topo-ML (Figure 9), we simply use XGBoost, MLP, and Transformer classifiers with the induced Betti vectors across all datasets. In the case of XGBoost, we adopted parametric tuning strategies. This approach involved configuring the learning rate to 0.1 and limiting the maximum depth to 5. As for the MLP architecture, it comprised a single input layer followed by two hidden layers, each containing 256, and 128 neurons respectively, all utilizing the RELU activation function. We also included an output layer in the architecture. The training of MLP models encompassed 100 epochs, employing a batch size of 64 for all datasets. We used Adam optimizer and retained the default settings for the remaining parameters. For Transformers, we used the same hyperparameter setting with the transformer encoder in Topo-VT model.

#### CNN Backbones

In our study, we investigated the alignment between topological features and current DL models using the architectures outlined in Section 3.2. Our rationale stems from the notion that while topological features capture global patterns within images, convolutional vectors focus more on local patterns. For the evaluation of our Topo-CNN models, we selected well-known CNN backbones, ResNet50, ResNet101 [34], Efficient-NetB0 [72], InceptionV3 [71], InceptionResNetv2 [71], MobileNetV2 [37], DenseNet121 [38], or VGG16 [63]. For Topo-VT model, we used SwinV2 [46].

#### Topo-CNN Hyperparameters

Our Topo-CNN model (Figure 3) integrates a pre-trained CNN with additional frozen layers, enhanced by a 1D Multi-Layer Perceptron (MLP). The augmented CNN includes a convolutional layer with 64 filters of size (3, 3) and ReLU activation, followed by a max-pooling layer with a pool size of (2, 2), a flattening layer, and a dense layer with 64 neurons. Simultaneously, the 1D MLP contributes with three dense layers having 800, 256, and 128 neurons respectively, all utilizing the ReLU activation function. The concatenated outputs from both models are further processed by fully connected layers, including 256, 128, and 128 neurons each with ReLU activation, culminating in the model’s final output. To investigate the utility of different vectorizations for topological features, we also employed Persistence Images (PI), and replaced Betti vectors with PI vectors. We give the details of this model (PI-CNN) in Appendix A.5.

#### Topo-VT Hyperparameters

Our transformer architecture (Figure 4) uses the pre-trained SwinV2 model [46] with all layers unfrozen for end-to-end learning. We extract feature maps from each Swin stage, compute Betti vector encodings 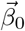 and 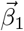, project them onto stage features, and apply cross-attention. The enriched features are then fed into the Swin head for baseline comparison. The model was trained for 100 epochs and using the AdamW optimizer with learning rate 1e-4, weight decay of 0.9, and dropout rate of 0.1. As previously stated, both Topo-Swin and the Betti encoders were trained together. In the Betti Transformer Encoder, each Betti vector has its own trainable encoder. Although Betti vectors are sequential data, they cannot be encoded to be at a specific patch of the image, so we let the model learn what the best positional encodings are. We utilize a Transformer Encoder which projects the Betti vectors to dimension 512. The encoders were trained with 4 heads, 4 layers, with a feed-forward dimension of 256, a dropout rate of 0.1, and ReLU activation. The encoders are trained end-to-end along with the Topo-Swin.

#### Runtimes

The time taken to generate topological features for the BUS-BRA dataset (consisting of 1875 images sized 275×375) on a personal laptop was 15 minutes for Betti vectors and 2.5 hours for persistence images. These computations were conducted on a personal laptop equipped with a 12th Gen Intel(R) Core(TM) i7 12650H processor clocked at 2.30 GHz, and supported by 64.0 GB of RAM. After the extraction of topological features, the overall average runtime for the Topo-CNN models was less than 3 hours on the same laptop.

#### Performance Metrics

We primarily used accuracy and ROC-AUC to evaluate model performance, but also report additional metrics, including F1 score, precision, and recall, in Appendix A.3.

### 4.3. Results

In this part, we discuss the results of our models utilizing topological features in different ways.

#### Topo-ML Model

As previously mentioned, the Topo-ML model (Figure 9) shows the standalone performance of topological vectors. Its primary advantages lie in its interpretability and computational efficiency. While its performance may not excel across all datasets, this straight-forward model offers a viable solution, particularly when data scarcity hampers the training of more complex CNN models. We present our Topo-ML results with different classifiers in Table 9. The performance of Topo-ML often aligns with several vanilla-CNN models in terms of accuracy. However, in terms of AUC, the Topo-ML model generally lags behind CNN models.

#### Topo-DL Models

In these models, our primary aim is to assess how topological features impact the enhancement of pretrained CNN and VT model performance. Our hypothesis posits that while DL models excel at capturing local image features, TDA grasps global features, offering complementary insights to DL models. Our findings, as depicted in Tables 2 and 3, clearly support this hypothesis.

**Table 3.**
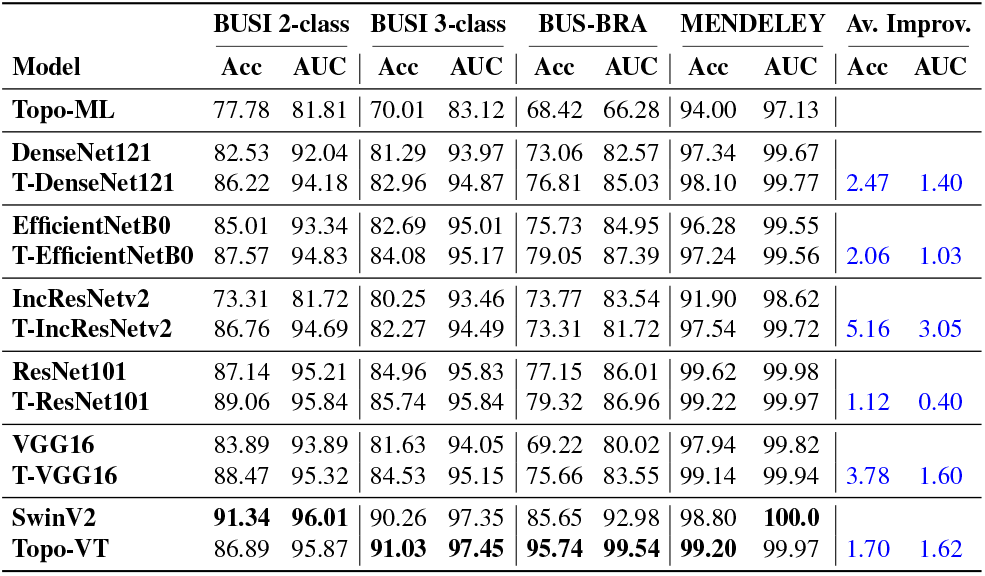
DL vs. Topo-DL. Performance comparison of vanilla DL models vs. Topo-DL models for breast ultrasound datasets. The last two columns show average improvements in accuracy and AUC. For each dataset, highest performances are given in **bold**.

Table 2 presents the results of our Topo-CNN model, showing that topological features consistently improve CNN performance across all datasets and backbone architectures. Incorporating topological features yields consistent accuracy and AUC gains of up to 6% for several backbones. Additionally, F1-scores for these models are reported in Table 11.

Table 3 summarizes the performance across all models, with the rightmost columns highlighting the average improvements contributed by topological features to pretrained deep learning models. We observe up to 5% and 3% average gains in accuracy and AUC, respectively. Given that these models are already pre-trained, these results indicate that topological features provide additional, valuable information that enhances the performance of these highly optimized models. Notably, the state-of-the-art VT model outperforms all CNN models in this setting, yet topological features still significantly boost the performance of these advanced architectures. Furthermore, our Topo-VT model outperforms recent Topo-DL approaches (Appendix A.2).

These results underscore the considerable potential for performance enhancement through the fusion of topological and DL features, even with a basic approach to inject topological features in CNN models. With tailored modifications based on specific domains, these findings suggest significant avenues for further performance gains leveraging the synergy between topological and DL features.

#### Limited Data Setting

We evaluated the effectiveness of topological features in limited data settings by randomly splitting the datasets into an 80:20 train-test ratio. With the test set held constant, we reduced the training set to 10, 20, and 50 images per class to evaluate performance with varying levels of limited data.

Our results in Table 4 show that topological features significantly enhance DL model performance, especially when training data is limited. These findings demonstrate the model’s robustness in low-data settings. Unlike traditional DL approaches, which often rely heavily on large datasets, topological features embed images directly into a latent space, reducing dependence on training set size.

**Table 4.**
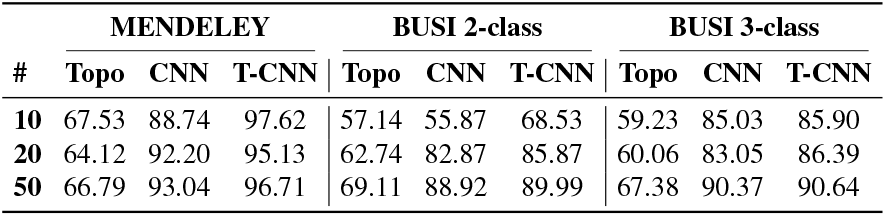
Performance with Limited Data: AUC scores for three models—TOPO (Betti vectors only), CNN (ResNet50), and TOPO-CNN (combined approach)—evaluated on the same test set with training data limited to 10, 20, or 50 samples per class.

| # | MENDELEY |  |  | BUSI 2-class |  |  | BUSI 3-class |  |  |
| --- | --- | --- | --- | --- | --- | --- | --- | --- | --- |
|  | Topo | CNN | T-CNN | Topo | CNN | T-CNN | Topo | CNN | T-CNN |
| 10 | 67.53 | 88.74 | 97.62 | 57.14 | 55.87 | 68.53 | 59.23 | 85.03 | 85.90 |
| 20 | 64.12 | 92.20 | 95.13 | 62.74 | 82.87 | 85.87 | 60.06 | 83.05 | 86.39 |
| 50 | 66.79 | 93.04 | 96.71 | 69.11 | 88.92 | 89.99 | 67.38 | 90.37 | 90.64 |

In few-shot learning or data-scarce conditions, our results indicate that topological features can improve model accuracy by providing reliable representations and stable anchors for each class, emphasizing their practical value in medical imaging, where labeled data is often scarce.

#### Topo-CNN in other medical domains

While we mainly focus on the breast ultrasound setting to maintain a clear scope, we also demonstrate the versatility and adaptability of our topological vectors and Topo-CNN model by showcasing their performance across various medical image formats. Utilizing the MedMNIST dataset [78], we employed three benchmark datasets RetinaMNIST (Fundus images), PneuMNIST (Chest X-ray) and BloodMNIST (Blood Cell) (see Table 8), applying our Betti-CNN model to 128 × 128 images with predefined splits. Notably, we observe up to 15% accuracy and 23% AUC improvements on CNN models by integrating topological features (Table 5).

**Table 5.** Topo-CNN on additional datasets. Performance comparison between vanilla-CNN and Topo-CNN models on MedMNIST datasets. For each dataset, the best two accuracy improvements are **underlined**, and the best two AUC improvements are marked **blue**.

| CNN Model | RetinaMNIST |  |  |  | PneuMNIST |  |  |  | BloodMNIST |  |  |  |
| --- | --- | --- | --- | --- | --- | --- | --- | --- | --- | --- | --- | --- |
|  | CNN |  | B-CNN |  | CNN |  | B-CNN |  | CNN |  | B-CNN |  |
|  | ACC | AUC | ACC | AUC | ACC | AUC | ACC | AUC | ACC | AUC | ACC | AUC |
| ResNet50 | 43.52 | 70.97 | 44.70 | 75.33 | **68.58** | 74.70 | **76.92** | 83.78 | 67.98 | **86.93** | 76.21 | **96.44** |
| EfficientNetB0 | 42.40 | 72.09 | 45.25 | 74.85 | 62.49 | **62.50** | 77.89 | **85.10** | 70.60 | 87.75 | 75.59 | 96.11 |
| InceptionV3 | 43.51 | 70.29 | 45.50 | 74.67 | 66.92 | 68.67 | 74.36 | 83.19 | **66.93** | 87.57 | **77.46** | 96.67 |
| MobileNetV2 | 43.00 | **68.02** | 45.75 | **73.71** | **62.66** | 65.18 | **77.24** | 83.91 | 69.85 | 87.41 | 77.76 | 96.44 |
| DenseNet121 | 43.50 | **70.38** | 45.00 | **75.18** | 63.57 | **60.01** | 77.08 | **82.01** | 71.85 | 87.62 | 77.02 | 96.42 |
| ResNet101 | **43.60** | 72.79 | **47.25** | 75.10 | 72.48 | 81.58 | 78.21 | 85.18 | 72.92 | **86.93** | 76.94 | **96.46** |
| VGG16 | **43.56** | 71.93 | **47.25** | 75.34 | 69.24 | 76.85 | 76.92 | 84.18 | **66.88** | 87.62 | **76.24** | 96.37 |

#### Complementary Nature of Topo & CNN models

Our results confirm that topological vectors improve DL performance. To validate their complementarity, we ran an 80:20 XGBoost experiment using 200-dim topological and 128-dim ResNet50 vectors (after 100 epochs). AUC and accuracy in Table 6 show the hybrid model outperforms both. The last two columns show the number of selected features in the hybrid model, with a balanced split further supporting complementarity.

**Table 6.**
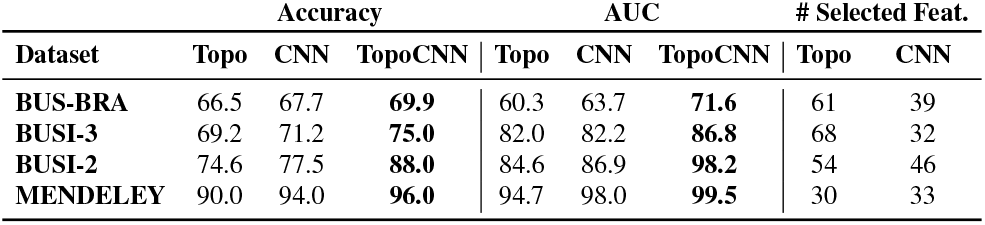
Complementarity of Topo and CNN Features. XGBoost performance on topological and CNN vectors, showing the hybrid model outperforms each alone. The balanced feature selection in the last two columns supports their complementarity.

| Dataset | Accuracy |  |  | AUC |  |  | # Selected Feat. |  |
| --- | --- | --- | --- | --- | --- | --- | --- | --- |
|  | Topo | CNN | TopoCNN | Topo | CNN | TopoCNN | Topo | CNN |
| BUS-BRA | 66.5 | 67.7 | **69.9** | 60.3 | 63.7 | **71.6** | 61 | 39 |
| BUSI-3 | 69.2 | 71.2 | **75.0** | 82.0 | 82.2 | **86.8** | 68 | 32 |
| BUSI-2 | 74.6 | 77.5 | **88.0** | 84.6 | 86.9 | **98.2** | 54 | 46 |
| MENDELEY | 90.0 | 94.0 | **96.0** | 94.7 | 98.0 | **99.5** | 30 | 33 |

**Table 7.** Performances of Topo-ML model with sublevel and superlevel filtrations are virtually same.

| Dataset | Class | Superlevel |  | Sublevel |  |
| --- | --- | --- | --- | --- | --- |
|  |  | Acc | AUC | Acc | AUC |
| BUSI | 2 | 76.63 | 81.12 | 77.78 | 81.81 |
| BUSI | 3 | 71.15 | 83.71 | 70.01 | 83.12 |
| MENDELEY | 2 | 94.57 | 98.04 | 94.00 | 97.13 |
| BUS-BRA | 2 | 67.65 | 66.40 | 68.42 | 66.28 |

**Table 8.** MedMNIST Dataset Details. * means varying image sizes.

| MedMNIST2D | Data Modality | Image size | # Classes | # Samples | Train/Valid/Test |
| --- | --- | --- | --- | --- | --- |
| PneuMNIST | Chest X-ray | 1650 × 1420* | 2 | 5,856 | 4,708 / 524 / 624 |
| RetinaMNIST | Fundus Camera | 1736 × 1824 | 5 | 1,600 | 1,080 / 120 / 400 |
| BreastMNIST | Breast Ultrasound | 500 × 500 | 2 | 780 | 546 / 78 / 156 |
| BloodMNIST | Blood Cell Microscope | 200 × 200 | 8 | 17,092 | 11,959 / 1,712 / 3,421 |

**Table 9.** ML Classifiers. Performance of our basic Topo-ML model with XGBoost, MLP and Transformer classifiers on breast ultrasound datasets.

|  |  | Accuracy |  |  | AUC |  |  |
| --- | --- | --- | --- | --- | --- | --- | --- |
| Dataset | Class | XGB | MLP | TF | XGB | MLP | TF |
| BUSI | 2 | 77.78 | **82.81** | 68.01 | 81.81 | **85.33** | 67.94 |
| BUSI | 3 | **70.01** | 66.28 | 60.38 | **83.12** | 78.29 | 73.86 |
| MENDELEY | 2 | 94.00 | **96.00** | 77.20 | 97.13 | **97.67** | 83.72 |
| BUS-BRA | 2 | 68.42 | 63.10 | **88.48** | 66.28 | 57.80 | **95.14** |

#### Alexander Duality

To empirically verify our Alexander duality result, we give performances of our Topo-ML model with topological vectors obtained via sublevel and superlevel filtrations. As claimed, sublevel and superlevel vectors are virtually mirror images of each other in complementary dimensions (Figure 5), and using one of these settings do not have any significant effect on the performance.

#### Statistical Analysis

To ensure a robust evaluation of model performance, we utilized 5 or 10-fold cross-validation, which inherently introduces variability in the performance metrics (e.g., accuracy, AUC, and F1-score). To further investigate the hypothesis that TDA can provide insights that complement CNN models, we further implemented a rigorous statistical analysis. This involved repeating the cross-validation process 20 times for each dataset across each model architecture, including B-CNN and PI-CNN. We then conducted a one-tailed paired t-test to statistically test whether the performance improvements observed with B-CNN or PI-CNN were significant over the baseline CNN model or if they could be attributed to random variation. Predicated on the premise that B-CNN generally outperforms P-CNN, we subjected this hypothesis to the same statistical test. To mitigate potential bias in our hypothesis testing, we complemented the t-test with the non-parametric Wilcoxon signed-rank test, a more robust test that does not assume a specific data distribution. The resulting *p*-values, which signify statistical significance, are reported in Table 13. Further details can be found in Appendix A.4, which provides statistical evidence that Topo-CNN models outperforms the baseline CNN models.

### 4.4. Interpretability of Topological Features

In this section, we explore the interpretability of topological features, focusing on Betti vectors as a straightforward yet effective method for vectorizing persistence diagrams, known for their high interpretability. In Figure 7, we present the Betti curves for the MENDELEY dataset, incorporating nonparametric confidence bands [27]. The figure reveals a distinct difference between the benign and malignant classes, particularly around *β*_0_(40) and *β*_1_(50).

To interpret these features: *β*_0_(40) refers to the number of components in the binary image χ_100_, where 100 represents the grayscale value corresponding to the 40^th^ thresh-old of the 100 thresholds within the [0, 255] grayscale range. Likewise, *β*_1_(50) denotes the count of white spots (holes) in χ_128_, with 128 corresponding to the grayscale value at the 50^th^ threshold.

In Fig. 6, we show boxplots and violin plots for these values, {*β*_0_(40)} and {*β*_1_(50)}, highlighting the clear distinction between the malignant and benign classes. Also, in Fig. 8, we display sample binary images χ_128_ with the 128 grayscale threshold for both classes, where the difference in the count of white spots (i.e., *β*_1_(50)) is clearly visible.

**Figure 6.**
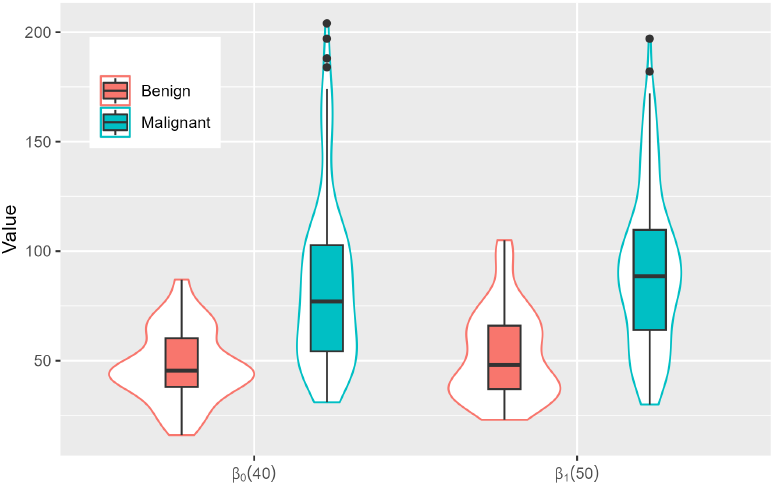
Boxplots with violin plots comparing the distributions of *β*_0_ (40) and *β*_1_ (50) coefficients of benign and malignant classes in MENDELEY dataset.

**Figure 7.**
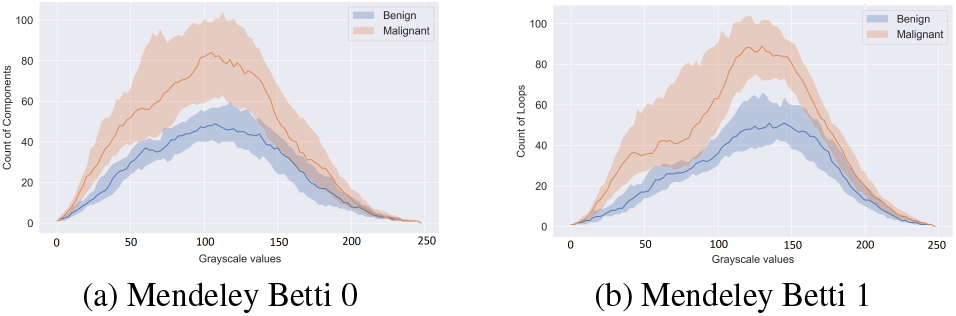
40% confidence bands for Betti-0 and Betti-1 curves for Benign and Malignant classes in the MENDELEY dataset. Bold curves represent the median for each class.

**Figure 8.**
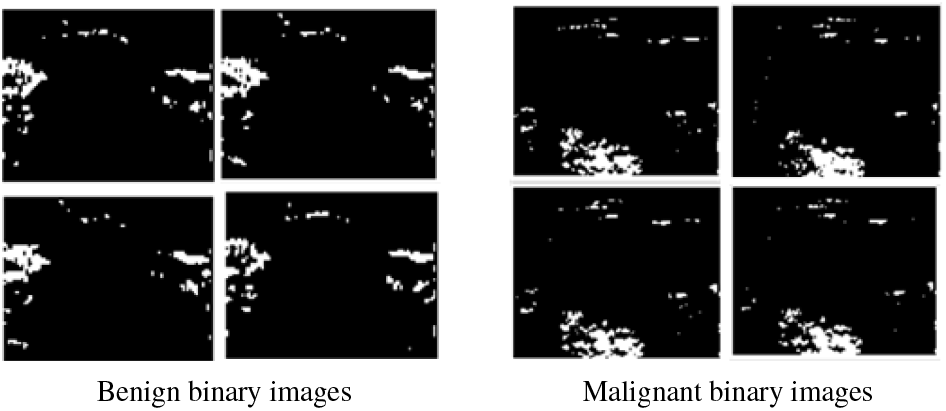
Sample binary images extracted from ultrasound images in the MENDELEY dataset. These binary images {χ_128_} are generated by marking all pixels with grayscale value ≤ 128 (threshold 50/100) black (Section 3.1). As suggested by Figures 6 and 7, the malignant images exhibit a significantly higher number of white spots (holes) compared to the benign images.

**Figure 9.**
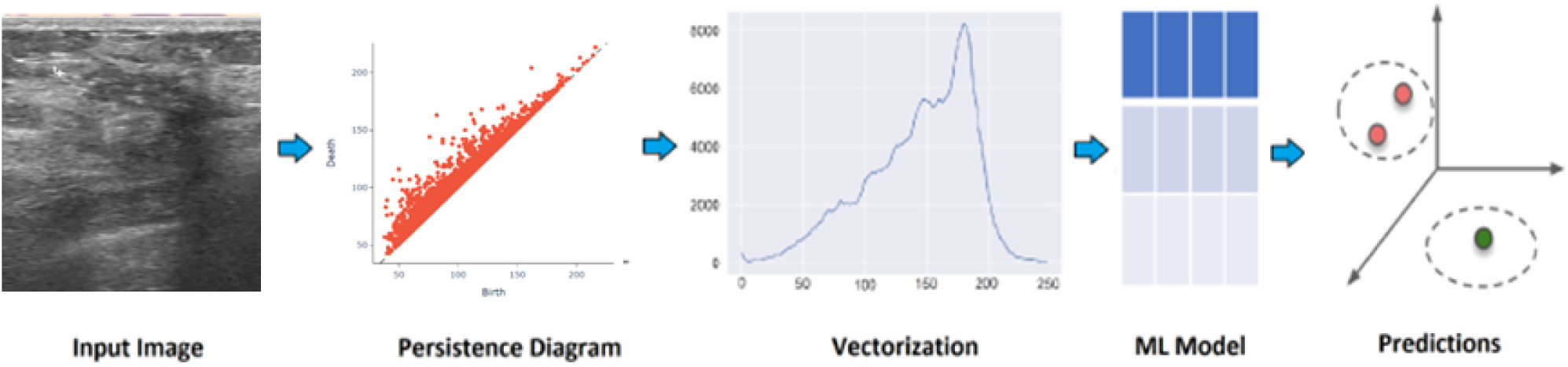
Topo-ML model. In our basic model, we first generate persistence diagrams for any input image. Next, we derive our topological feature vectors, which are then inputted into an ML classifier to produce classification results.

## 5. Conclusion

We investigated the effectiveness of topological features in enhancing DL models for breast cancer screening. Notably, we found that TDA offers a distinct approach to image analysis, diverging from conventional DL models and addressing key limitations. Our findings yield two main results. First, when combined with simple ML methods, topological features offer a viable alternative to DL models due to their efficiency, interpretability, and solid performance on limited data. Second, when integrated with DL models, they significantly improve accuracy and robustness. Moving forward, we aim to leverage this potential to develop specialized tools for medical image analysis by merging TDA outputs with DL models to create advanced, reliable decision support systems for breast cancer screening and beyond.

## Data Availability

All datasets are publicly available and links are provided in the paper.

https://www.kaggle.com/datasets/subhajournal/busi-breast-ultrasound-images-dataset

## Acknowledgements

This work was partially supported by National Science Foundation under grants DMS-2113674, DMS-2210912, DMS-2220613, and DMS-2229417. The authors acknowledge the Texas Advanced Computing Center (TACC) at UT Austin for providing computational resources that have contributed to the research results reported within this paper. http://www.tacc.utexas.edu.

## Appendix

### A. More on Experimental Results

In this part, we give additional results and details of our models.

#### A.1. MedMNIST datasets

While the main part of our paper focuses on the Breast Ultrasound setting to maintain a clear scope, we also demonstrate the versatility and adaptability of our topological vectors and their significant contributions to existing deep learning models by showcasing their performance across various medical image formats. Utilizing the MedMNIST dataset [78], we employed three benchmark datasets, RETINAMNIST, PneuMNIST and BloodMNIST(see Table 8), applying our Betti-CNN model to 128 *×* 128 images with predefined splits. Note that the BreastMNIST dataset corresponds to the BUSI dataset used in this paper. The performance of our model for BUSI is detailed in the main text (see Table 2).

#### A.2. Comparison with Other Topo-DL Models

We conducted experiments using PHG-Net [50], and the performance comparison in terms of AUC and Accuracy is presented in Table 10.

**Table 10.** Comparison of Topo-DL Models. AUC and Accuracy comparisons for PHG-Net and Topo-Swin.

| Dataset | Accuracy |  | AUC |  |
| --- | --- | --- | --- | --- |
|  | PHG-NET | Topo-VT | PHG-NET | Topo-VT |
| BUSI-2 | 86.6 | **86.9** | 94.7 | **95.9** |
| BUSI-3 | 87.4 | **91.0** | 96.7 | **97.5** |
| BUS-BRA | 83.0 | **95.7** | 90.1 | **99.5** |
| MENDELEY | **99.6** | 99.2 | 99.6 | **99.9** |

**Table 11.** Comparison of F1-scores between CNN and Topo-CNN models.

| MODEL | MENDELEY |  |  | BUS-BRA |  |  | BUSI 2-Labels |  |  | BUSI 3-Labels |  |  |
| --- | --- | --- | --- | --- | --- | --- | --- | --- | --- | --- | --- | --- |
|  | CNN | B-CNN | P-CNN | CNN | B-CNN | P-CNN | CNN | B-CNN | P-CNN | CNN | B-CNN | P-CNN |
| DenseNet121 | 97.72 | 98.33 | 99.81 | 74.65 | 77.37 | 76.32 | 84.32 | 87.09 | 88.31 | 81.29 | 83.47 | 84.63 |
| EfficientNetB0 | 96.79 | 97.51 | 97.60 | 76.88 | 79.87 | 78.20 | 85.86 | 88.41 | 87.59 | 82.69 | 84.42 | 84.49 |
| IncResNetV2 | 93.18 | 97.86 | 99.42 | 75.25 | 74.35 | 75.16 | 74.35 | 87.59 | 87.73 | 80.25 | 82.77 | 83.64 |
| InceptionV3 | 98.79 | 98.66 | 100.0 | 73.62 | 73.30 | 73.07 | 84.08 | 85.85 | 86.22 | 79.77 | 82.30 | 81.16 |
| MobileNetV2 | 98.66 | 98.35 | 99.63 | 75.25 | 76.78 | 76.51 | 85.35 | 86.94 | 87.28 | 82.65 | 84.01 | 80.56 |
| ResNet50 | 99.77 | 99.24 | 99.62 | 77.80 | 80.04 | 78.94 | 86.97 | 88.58 | 88.20 | 83.87 | 85.18 | 85.33 |
| ResNet101 | 99.73 | 99.36 | 98.91 | 78.62 | 80.03 | 79.91 | 88.42 | 89.86 | 88.98 | 84.96 | 85.98 | 86.02 |
| VGG16 | 98.55 | 99.27 | 99.23 | 72.30 | 76.35 | 74.68 | 86.37 | 89.28 | 89.86 | 81.63 | 84.86 | 84.57 |

**Table 12.** Performance metrics for SwinV2 and Topo-VT models.

| Dataset | SwinV2 |  |  |  |  | Topo-VT |  |  |  |  |
| --- | --- | --- | --- | --- | --- | --- | --- | --- | --- | --- |
|  | Acc | AUC | Prec | Recall | F1 | Acc | AUC | Prec | Recall | F1 |
| BUSI-2 | 91.34 | 96.01 | 91.29 | 89.14 | 89.84 | 86.89 | 95.87 | 89.79 | 81.00 | 83.10 |
| BUSI-3 | 90.26 | 97.35 | 90.84 | 88.11 | 89.01 | 91.03 | 97.45 | 90.05 | 90.62 | 90.13 |
| BUS-BRA | 85.65 | 92.98 | 84.38 | 83.68 | 83.42 | 95.74 | 99.54 | 94.83 | 95.57 | 95.19 |
| MENDELEY | 98.80 | 100.0 | 99.09 | 98.50 | 98.71 | 99.20 | 99.97 | 99.38 | 99.00 | 99.15 |

#### A.3. Additional Performance Metrics

In medical image analysis, F1-scores is a very common metric incorporating both precision and recall. Precision (also called positive predictive value) measures the accuracy of positive predictions, while recall (also called sensitivity or true positive rate) measures the proportion of actual positives that were correctly identified by the model. The F1-score is the harmonic mean of precision and recall and is calculated as 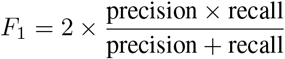.

In Table 11, we present the F1-scores of our models. It’s worth noting that although the improvements may appear marginal, our findings in Table 13 demonstrate that most of these enhancements are statistically highly significant.

#### A.4. Significance Test

In Table 2 and Table 11, we outlined the three performance metrics for vanilla-CNN and our Topo-CNN models, highlighting the integration of topological features into conventional CNN architectures has potential to enhance the breast cancer screening. To evaluate the statistical significance of these enhancements, we utilized two distinct methods: the t-test and the Wilcoxon signed-rank test. The results, including all *p*-values, are documented in Table 13 and Table 14. A significant majority of these *p*-values fell below the 0.05 threshold, indicating statistical significance, and are accordingly highlighted in blue for easy identification.

**Table 13.** Summary of *p*-values derived from t-tests indicating the performance disparity between vanilla-CNN and Topo-CNN models (Betti-CNN and Persistence Image-CNN) through t-test for BUSI, BUS-BRA, and MENDELEY datasets. To condense, we denote *e*^*n*^ as 10^*n*^. Outcomes with *p <* 0.05 are statistically significant and are highlighted in blue.

| BUSI 2-label |  |  |  |  |  |  |  |  |  |
| --- | --- | --- | --- | --- | --- | --- | --- | --- | --- |
| MODEL | CNN vs B-CNN |  |  | CNN vs P-CNN |  |  | P-CNN vs B-CNN |  |  |
|  | ACC | AUC | F-1 | ACC | AUC | F-1 | ACC | AUC | F-1 |
| DenseNet121 | 8.5e<sup>-09</sup> | 1.6e<sup>-09</sup> | 7.6e<sup>-08</sup> | 2.1e<sup>-05</sup> | 6.4e<sup>-03</sup> | 2.7e<sup>-04</sup> | 9.4e<sup>-01</sup> | 2.1e<sup>-01</sup> | 8.7e<sup>-01</sup> |
| EfficientNetB0 | 5.6e<sup>-09</sup> | 1.8e<sup>-12</sup> | 2.2e<sup>-10</sup> | 3.0e<sup>-03</sup> | 6.8e<sup>-01</sup> | 1.9e<sup>-02</sup> | 4.3e<sup>-01</sup> | 9.1e<sup>-03</sup> | 1.7e<sup>-01</sup> |
| InceptionV3 | 1.5e<sup>-06</sup> | 1.1e<sup>-08</sup> | 1.3e<sup>-05</sup> | 4.5e<sup>-02</sup> | 1.5e<sup>-01</sup> | 7.2e<sup>-02</sup> | 6.4e<sup>-01</sup> | 1.1e<sup>-01</sup> | 6.0e<sup>-01</sup> |
| IncResNetV2 | 1.1e<sup>-14</sup> | 1.4e<sup>-15</sup> | 6.3e<sup>-15</sup> | 2.9e<sup>-12</sup> | 1.0e<sup>-12</sup> | 2.4e<sup>-12</sup> | 8.0e<sup>-01</sup> | 1.4e<sup>-01</sup> | 5.8e<sup>-01</sup> |
| MobileNetV2 | 4.3e<sup>-06</sup> | 3.6e<sup>-02</sup> | 3.3e<sup>-05</sup> | 1.4e<sup>-03</sup> | 9.2e<sup>-01</sup> | 2.0e<sup>-02</sup> | 8.4e<sup>-01</sup> | 5.1e<sup>-02</sup> | 6.4e<sup>-01</sup> |
| ResNet101 | 1.4e<sup>-07</sup> | 5.5e<sup>-05</sup> | 6.2e<sup>-06</sup> | 7.6e<sup>-02</sup> | 7.6e<sup>-01</sup> | 2.9e<sup>-01</sup> | 3.9e<sup>-01</sup> | 7.0e<sup>-02</sup> | 2.1e<sup>-01</sup> |
| ResNet50 | 5.9e<sup>-10</sup> | 8.9e<sup>-07</sup> | 3.2e<sup>-08</sup> | 3.0e<sup>-03</sup> | 6.3e<sup>-01</sup> | 6.7e<sup>-02</sup> | 6.2e<sup>-01</sup> | 4.6e<sup>-02</sup> | 3.2e<sup>-01</sup> |
| VGG16 | 9.4e<sup>-12</sup> | 5.5e<sup>-11</sup> | 7.3e<sup>-11</sup> | 1.2e<sup>-06</sup> | 2.8e<sup>-02</sup> | 3.1e<sup>-04</sup> | 9.1e<sup>-01</sup> | 2.9e<sup>-01</sup> | 7.5e<sup>-01</sup> |

| BUSI 3-label |  |  |  |  |  |  |  |  |  |
| --- | --- | --- | --- | --- | --- | --- | --- | --- | --- |
| MODEL | CNN vs B-CNN |  |  | CNN vs P-CNN |  |  | P-CNN vs B-CNN |  |  |
|  | ACC | AUC | F-1 | ACC | AUC | F-1 | ACC | AUC | F-1 |
| DenseNet121 | 9.8e<sup>-09</sup> | 1.6e<sup>-07</sup> | 7.3e<sup>-10</sup> | 1.2e<sup>-10</sup> | 2.4e<sup>-01</sup> | 1.2e<sup>-11</sup> | 1.0e<sup>-00</sup> | 1.1e<sup>-05</sup> | 1.0e<sup>-00</sup> |
| EfficientNetB0 | 1.6e<sup>-04</sup> | 1.1e<sup>-01</sup> | 1.6e<sup>-05</sup> | 5.9e<sup>-05</sup> | 1.0e<sup>-00</sup> | 2.1e<sup>-06</sup> | 5.2e<sup>-01</sup> | 1.4e<sup>-06</sup> | 5.9e<sup>-01</sup> |
| InceptionV3 | 2.6e<sup>-05</sup> | 7.1e<sup>-06</sup> | 2.6e<sup>-06</sup> | 1.3e<sup>-02</sup> | 1.0e<sup>-00</sup> | 6.0e<sup>-04</sup> | 1.0e<sup>-02</sup> | 2.5e<sup>-08</sup> | 6.9e<sup>-03</sup> |
| IncResNetV2 | 2.2e<sup>-04</sup> | 9.5e<sup>-07</sup> | 1.6e<sup>-05</sup> | 2.1e<sup>-07</sup> | 1.9e<sup>-01</sup> | 1.2e<sup>-08</sup> | 9.9e<sup>-01</sup> | 2.1e<sup>-06</sup> | 9.9e<sup>-01</sup> |
| MobileNetV2 | 1.4e<sup>-03</sup> | 2.7e<sup>-01</sup> | 8.1e<sup>-05</sup> | 1.0e<sup>-00</sup> | 1.0e<sup>-00</sup> | 1.0e<sup>-00</sup> | 2.4e<sup>-05</sup> | 8.9e<sup>-11</sup> | 1.4e<sup>-05</sup> |
| ResNet101 | 3.8e<sup>-02</sup> | 4.9e<sup>-01</sup> | 1.3e<sup>-02</sup> | 6.0e<sup>-03</sup> | 1.0e<sup>-00</sup> | 8.5e<sup>-05</sup> | 4.4e<sup>-01</sup> | 2.0e<sup>-05</sup> | 5.5e<sup>-01</sup> |
| ResNet50 | 6.2e<sup>-04</sup> | 7.5e<sup>-01</sup> | 1.5e<sup>-04</sup> | 1.0e<sup>-03</sup> | 1.0e<sup>-00</sup> | 8.6e<sup>-05</sup> | 6.1e<sup>-01</sup> | 1.3e<sup>-08</sup> | 7.0e<sup>-01</sup> |
| VGG16 | 6.6e<sup>-08</sup> | 2.8e<sup>-07</sup> | 1.1e<sup>-08</sup> | 5.9e<sup>-08</sup> | 7.1e<sup>-01</sup> | 4.0e<sup>-09</sup> | 2.7e<sup>-01</sup> | 1.5e<sup>-07</sup> | 2.5e<sup>-01</sup> |

| BUS-BRA |  |  |  |  |  |  |  |  |  |
| --- | --- | --- | --- | --- | --- | --- | --- | --- | --- |
| MODEL | CNN vs B-CNN |  |  | CNN vs P-CNN |  |  | P-CNN vs B-CNN |  |  |
|  | ACC | AUC | F-1 | ACC | AUC | F-1 | ACC | AUC | F-1 |
| DenseNet121 | 4.0e<sup>-08</sup> | 2.8e<sup>-06</sup> | 6.4e<sup>-08</sup> | 3.8e<sup>-03</sup> | 9.7e<sup>-01</sup> | 1.1e<sup>-02</sup> | 1.2e<sup>-01</sup> | 5.0e<sup>-05</sup> | 5.0e<sup>-02</sup> |
| EfficientNetB0 | 2.0e<sup>-11</sup> | 1.0e<sup>-09</sup> | 4.4e<sup>-11</sup> | 1.3e<sup>-04</sup> | 9.6e<sup>-01</sup> | 1.2e<sup>-03</sup> | 5.3e<sup>-02</sup> | 2.2e<sup>-07</sup> | 7.8e<sup>-04</sup> |
| InceptionV3 | 3.6e<sup>-01</sup> | 9.9e<sup>-01</sup> | 8.5e<sup>-01</sup> | 1.2e<sup>-01</sup> | 1.0e<sup>-00</sup> | 8.2e<sup>-01</sup> | 8.5e<sup>-01</sup> | 1.3e<sup>-06</sup> | 3.4e<sup>-01</sup> |
| IncResNetV2 | 8.0e<sup>-01</sup> | 1.0e<sup>-00</sup> | 9.5e<sup>-01</sup> | 1.2e<sup>-02</sup> | 1.0e<sup>-00</sup> | 5.7e<sup>-01</sup> | 9.8e<sup>-01</sup> | 3.5e<sup>-02</sup> | 8.8e<sup>-01</sup> |
| MobileNetV2 | 4.8e<sup>-07</sup> | 1.8e<sup>-06</sup> | 3.7e<sup>-05</sup> | 4.3e<sup>-06</sup> | 1.0e<sup>-00</sup> | 4.3e<sup>-03</sup> | 8.1e<sup>-01</sup> | 1.8e<sup>-06</sup> | 2.8e<sup>-01</sup> |
| ResNet101 | 3.8e<sup>-07</sup> | 6.5e<sup>-04</sup> | 1.0e<sup>-04</sup> | 1.7e<sup>-05</sup> | 9.6e<sup>-01</sup> | 8.6e<sup>-03</sup> | 8.7e<sup>-01</sup> | 1.1e<sup>-03</sup> | 4.1e<sup>-01</sup> |
| ResNet50 | 7.4e<sup>-05</sup> | 6.5e<sup>-04</sup> | 3.0e<sup>-04</sup> | 5.2e<sup>-03</sup> | 9.7e<sup>-01</sup> | 8.0e<sup>-02</sup> | 2.7e<sup>-01</sup> | 2.3e<sup>-04</sup> | 6.2e<sup>-02</sup> |
| VGG16 | 2.3e<sup>-17</sup> | 2.9e<sup>-11</sup> | 2.2e<sup>-13</sup> | 5.3e<sup>-11</sup> | 1.0e<sup>-00</sup> | 6.3e<sup>-06</sup> | 5.1e<sup>-02</sup> | 2.0e<sup>-09</sup> | 3.8e<sup>-04</sup> |

| MENDELEY |  |  |  |  |  |  |  |  |  |
| --- | --- | --- | --- | --- | --- | --- | --- | --- | --- |
| MODEL | CNN vs B-CNN |  |  | CNN vs P-CNN |  |  | P-CNN vs B-CNN |  |  |
|  | ACC | AUC | F-1 | ACC | AUC | F-1 | ACC | AUC | F-1 |
| DenseNet121 | 1.3e<sup>-03</sup> | 3.3e<sup>-02</sup> | 8.0e<sup>-04</sup> | 4.7e<sup>-09</sup> | 5.3e<sup>-07</sup> | 7.4e<sup>-09</sup> | 1.0e<sup>-00</sup> | 1.0e<sup>-00</sup> | 1.0e<sup>-00</sup> |
| EfficientNetB0 | 2.6e<sup>-03</sup> | 4.3e<sup>-01</sup> | 9.9e<sup>-03</sup> | 1.2e<sup>-01</sup> | 5.0e<sup>-06</sup> | 1.7e<sup>-01</sup> | 5.8e<sup>-01</sup> | 1.0e<sup>-00</sup> | 5.5e<sup>-01</sup> |
| InceptionV3 | 6.7e<sup>-01</sup> | 9.5e<sup>-01</sup> | 7.6e<sup>-01</sup> | 3.7e<sup>-09</sup> | 8.9e<sup>-05</sup> | 6.8e<sup>-09</sup> | 1.0e<sup>-00</sup> | 1.0e<sup>-00</sup> | 1.0e<sup>-00</sup> |
| IncResNetV2 | 1.1e<sup>-12</sup> | 2.8e<sup>-10</sup> | 4.5e<sup>-12</sup> | 7.1e<sup>-15</sup> | 7.7e<sup>-14</sup> | 4.0e<sup>-15</sup> | 1.0e<sup>-00</sup> | 1.0e<sup>-00</sup> | 1.0e<sup>-00</sup> |
| MobileNetV2 | 9.6e<sup>-01</sup> | 9.7e<sup>-01</sup> | 9.7e<sup>-01</sup> | 3.3e<sup>-03</sup> | 1.5e<sup>-06</sup> | 6.5e<sup>-03</sup> | 1.0e<sup>-00</sup> | 1.0e<sup>-00</sup> | 1.0e<sup>-00</sup> |
| ResNet101 | 1.0e<sup>-00</sup> | 8.5e<sup>-01</sup> | 1.0e<sup>-00</sup> | 8.6e<sup>-01</sup> | 7.3e<sup>-01</sup> | 8.9e<sup>-01</sup> | 2.9e<sup>-01</sup> | 3.1e<sup>-01</sup> | 2.5e<sup>-01</sup> |
| ResNet50 | 1.0e<sup>-00</sup> | 9.6e<sup>-01</sup> | 1.0e<sup>-00</sup> | 5.8e<sup>-01</sup> | 4.3e<sup>-02</sup> | 7.0e<sup>-01</sup> | 9.2e<sup>-01</sup> | 1.0e<sup>-00</sup> | 8.8e<sup>-01</sup> |
| VGG16 | 3.9e<sup>-07</sup> | 1.9e<sup>-03</sup> | 1.0e<sup>-05</sup> | 3.4e<sup>-03</sup> | 4.4e<sup>-06</sup> | 4.5e<sup>-02</sup> | 5.7e<sup>-01</sup> | 1.0e<sup>-00</sup> | 4.5e<sup>-01</sup> |

**Table 14.**
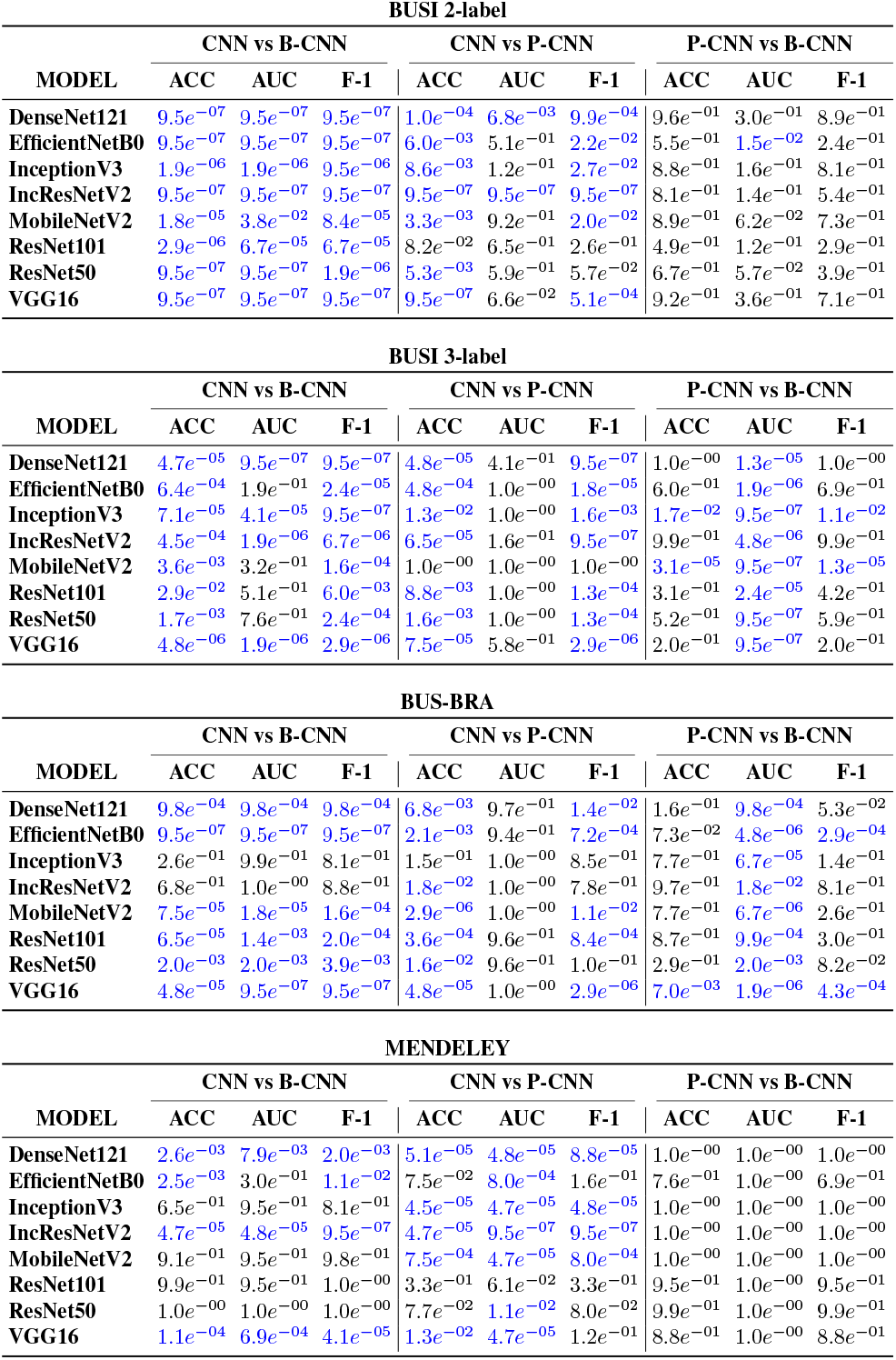
Summary of *p*-values derived from Wilcoxon signed-rank test indicating the performance disparity between vanilla-CNN and Topo-CNN models (Betti-CNN and Persistence Image-CNN) through t-test for BUSI, BUS-BRA, and MENDELEY datasets. To condense, we denote *e*^*n*^ as 10^*n*^. Outcomes with *p <* 0.05 are statistically significant and are highlighted in blue.

Our analysis showed that the B-CNN model outperformed the standard CNN model with a notable margin, as evidenced by the prevalence of blue in its respective subsection. Similarly, the P-CNN model demonstrated a significant improvement over the conventional CNN, with more than half of the comparative tests yielding significant results, also marked in blue. When comparing P-CNN against B-CNN, our findings suggest a superior performance by B-CNN, particularly on the BUSI and BUS-BRA datasets.

To conclude, the statistical evidence supports that both Topo-CNN models, namely P-CNN and B-CNN, significantly surpass the baseline CNN model in terms of performance by leveraging topological features. Despite this, the performance gap between P-CNN and B-CNN, while present, often does not reach statistical significance, suggesting a nuanced comparison that varies by dataset and context.

#### A.5. Persistence Images and PI-CNN model

To explore the effect of vectorization on our models, we also employed a different vectorization to replace Betti vectors in our models.

##### Persistence Images

Persistence Images is one of the most common vectorization methods in TDA, introduced by Adams et al. [2]. Unlike most vectorizations, Persistence Images, as the name suggests, produce 2*D*-arrays (tensors). The idea is to capture the location of the points in the PDs with a multivariable function by using the 2*D* Gaussian functions centered at these points. For *PD*(**G**) = {(*b*_*i*_, *d*_*i*_)}, let *ϕ*_*i*_ represent a 2*D*-Gaussian centered at the point (*b*_*i*_, *d*_*i*_) ∈ ℝ^2^. Then, one defines a multivariable function, *Persistence Surface*, 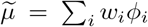 where *w*_*i*_ is the weight, mostly a function of the life span *d*_*i*_ − *b*_*i*_. To represent this multivariable function as a 2*D*-vector, one defines a *k × l* grid (resolution size) on the domain of 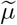, i.e., threshold domain of *PD*(**G**). Then, one obtains the *Persistence Image*, a 2*D*-vector (matrix) 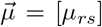 of size *k × l* such that

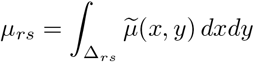

where Δ_*rs*_ = pixel with index *rs* in the *k × l* grid.

Note that the resolution size *k × l* is independent of the number of thresholds used in the filtering; the choice of *k* and *l* is completely up to the user. There are two other important tuning parameters for persistence images, namely the weight *w*_*i*_ and the variance *σ* (the width of the Gaussian functions). Like Silhouettes, one can choose *w*_*i*_ = (*d*_*i*_ *™ b*_*i*_)^*p*^ to emphasize large or small features in the PD. Similarly, the width parameter *σ* determines the sharpness of Gaussian, where smaller *σ* would make the Gaussian functions more like Dirac δ-function, and larger *σ* would make the Gaussians flat. Depending on the context, *σ* can be chosen a constant (e.g. *σ* = 0.1) or depending on the point (*b*_*i*_, *d*_*i*_), e.g., *σ*_*i*_ = *k*(*d*_*i*_ − *b*_*i*_) for some constant *k >* 0.

##### Hyperparameters for PI-CNN model

For each image χ, we first generated two 50 by 50 persistence images 𝒫ℐ_0_(χ) and 𝒫ℐ_1_(χ) corresponding to homology groups 0 and 1 respectively. These two persistence images were concatenated to obtain 50 by 100 matrix, which serves as a topological representation for each 2D image.

Our PI-CNN model (Figure Figure 3) integrates a pretrained CNN with inputs of size 224 × 224 with additional frozen layers, enhanced by a 2D Multi-Layer Perceptron (2D MLP). The augmented CNN includes a convolutional layer with 64 filters of size (3, 3) and ReLU activation, followed by a max-pooling layer with a pool size of (2, 2), a flatten layer, and a dense layer with 64 neurons. Simultaneously, the 2D MLP consists of a convolutional layer with 32 filters of size (3, 3), a max pooling layer of size (2, 2), a flatten layer, and a dense layer with 128 neurons using ReLU activation. The concatenated outputs from both models are further processed by fully connected layers, including 256, 128, and 128 neurons each with ReLU activation, culminating in the model’s final output.

### B. Alexander Duality for Cubical Persistence

In this section, we show the duality between sublevel filtration and superlevel filtration for cubical persistence. We note that in [9], the authors prove a duality result for cubical complexes, however their duality results are completely unrelated to ours. In their work, the authors study pixel connectivity in cubical complexes and construct *dual graph filtrations* for given cubical complex filtration. In our case, we relate one cubical complex filtration to another one (sublevel-superlevel) and show their duality in complementary dimensions.

First, we recall some earlier results from algebraic topology, which is essential for the proof. Next, we give the setup and introduce the notation which we use in upcoming sections.

#### B.1. Preliminaries

In this part, we recall some of the well-known results which we use in the proof of the duality theorem [33].

##### Lemma 2 (Alexander Duality)

*Let* χ *be a compact, locally contractible subspace of d-sphere* **S**^*d*^. *Then*,

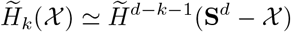

*where* 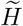 *represents* reduced *(co)homology*.

Since we only use field coefficients **F** in persistence modules, the Universal Coefficients Theorem [33] comes in a very simple form as follows.

##### Lemma 3 (Universal Coefficients Theorem)

*Let* **F** *be a field. Then, for any k*,

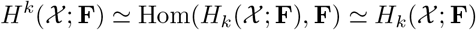

Next, we will need the following duality result between persistent homology and persistent cohomology [21].

##### Lemma 4 (Persistent Cohomology)

*For a given filtration* {χ_*n*_}, *persistent homology (H*_*k*_( χ_*i*_) → *H*_*k*_( χ_*j*_)*) and persistent cohomology (H*^*k*^( χ_*i*_) ← *H*^*k*^( χ_*j*_)*) have identical barcodes:*

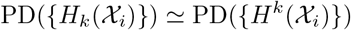

Finally, we quote the celebrated result on the uniqueness of the barcode decomposition in persistence module [10].

##### Lemma 5 (Krull-Schmidt Theorem)

*Any finite-dimensional persistence module has a unique (up to isomorphism) decomposition into indecomposable modules*.

In other words, the dimension of the homology groups {*H*_*k*_(χ_*i*_)}, and ranks of the homomorphisms *φ*_*ij*_: {*H*_*k*_(χ_*i*_) → *H*_*k*_(χ_*j*_)}, uniquely determines the persistence barcode. Here, each bar in the persistence barcode corresponds to an indecomposable module in the theorem.

#### B.2. Notation and Setup

In the following, we will work with 3 different cubical complexes (CC): The original CC χ, the extended CC 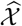, and the extended sphere 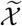.

**The image** χ : Let χ be a *d*-dimensional cubical complex of resolution 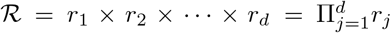 Let *η* = (*η*_1_, *η*_2_, …, *η*_*N*_) represent the index of the voxels in the cubical complex χ, i.e., 1 ≤ *η*_*j*_ *≤ r*_*j*_. By abusing the notation, we will call the collection of voxels in χ as ℛ = {Δ_*η*_}. Let *f* : ℛ → ℝ be the filtering function assigning each voxel Δ_*η*_ with corresponding (color) value *f* (*η*). Let *m* = min *f* and *M* = max *f*. Notice that the only interesting persistence diagrams for *d*-dimensional cubical complex are *k* = 0, 1, 2, …, *d ™* 1 as for *k ≥ d*, PD_*k*_(χ) is trivial.

**The extended (padded) image** 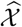: First, we induce a slightly larger complex 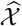 of resolution 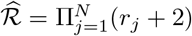 with indices *η* = (*η*_1_, *η*_2_, …, *η*_*N*_) again where 0 ≤ *η*_*j ≤*_ *r*_*j*_ + 1. In other words, we attach a padding of thickness-1 along the boundary of our original cubical complex χ. Then, we extend *f* to 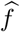 such that 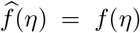 for any *η* ∈ *ℛ*, and 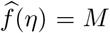 for any 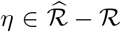, e.g., the value of 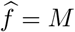 for any voxel in the boundary shell 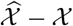.

**The extended sphere** 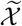: Our second extension is to double 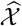 topologically and obtain a *d*-sphere 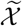. In particular, glueing two *d*-balls along their boundaries with an identity map gives a *d*-sphere [33]. Here, we do the same by taking an exact copy of 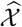 and glue it to the original one along the boundary. Hence, we double the number of voxels in 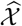, and we have a new index set 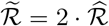. Again, we extend *f* to 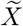 as before. i.e., 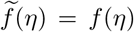 for any *η* ∈ *R*, and 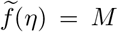 for any 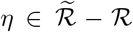. In other words, in the and *d*-sphere 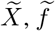 is defined as *M* for any voxel in 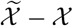.

In the following, we will represent *k*^*th*^ persistence diagram for sublevel filtration as 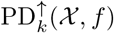 and *k*^*th*^ persistence diagram for superlevel filtration as 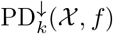.

##### Proof of the Theorem

Now, we are ready to prove our main duality result. In the following theorem, we prove the equivalence of persistence diagrams of sublevel and superlevel filtrations in complementary dimensions. In Remark 7, we give the explicit description of the correspondence between the bars in these persistence barcodes.

###### Theorem 6

*Let* χ *be a cubical complex of dimension d, and f be the filtering function on* χ. *Let* 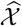 *be its extended image with* 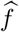. *Then, the persistence diagrams for the sub-level and superlevel filtration of* 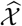 *with respect to* 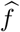 *in complementary dimensions are equivalent*.

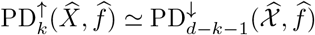

*Proof:* We prove the theorem in 2 steps. In the first step, we prove the duality in the sphere setting 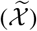. In this version, we prove the equivalence between the *reduced* persistence diagrams which means that for dimension *k* = 0, the main barcode corresponding to the connected component with infinite lifespan is removed. Then, in the second step, we adapt this result to *extended* images 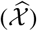 setting and finish the proof.

**Step 1:** Let 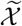 be the extended sphere of, and *f* be the corresponding extension of 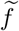. Then, the following *reduced* persistence diagrams are equivalent:

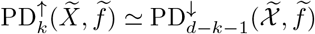

*Proof of Step 1:* Let *t*_0_ *< t*_1_ = min *f < t*_2_ *<* · · · *< t*_*N*−1_ *< t*_*N*_ = max *f* be the threshold set for *f*. Let 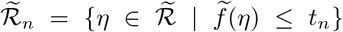. As before, define the sublevel filtration for 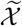 such that 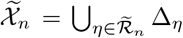. Similarly, let 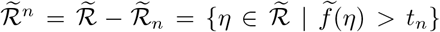. Now, the superlevel filtration can be defined as 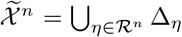, ie.,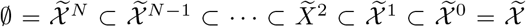. Note that to avoid crowding the notation, we slightly modify superlevel filtration with strict inequality 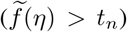. This only shifts the indexes by one from the original superlevel filtration 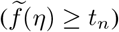 (Section 3.1).

By definition, we have 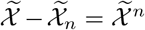. Recall that 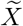 is a *d*-sphere. Hence, by Alexander duality (Lemma 2), for any 0 ≤ *k* ≤ *d* − 1, we have

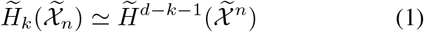

Notice that the Alexander duality uses reduced homology, hence infinite barcode for *k* = 0 is removed in this correspondence. Now, consider the persistent cohomology induced by the filtration 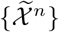, where 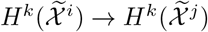 as 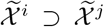 for *i < j*. Notice that by the naturality of *H*^*k*^( χ; **F**) = Hom(*H*_*k*_( χ; **F**), **F**) (Lemma 3), we have the following commutative diagram:

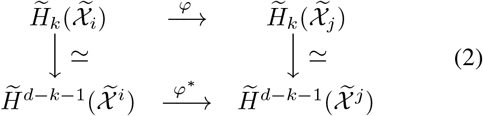

Now, we claim that *k*^*th*^ persistent homology module for 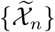 is equivalent to (*d* − *k* − 1)^*th*^ persistent cohomology module of 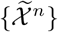. By Equation 1, we have the dimensions of the corresponding homologies are same 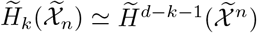. By Equation 2, we have ranks of the maps 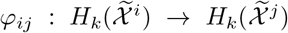 and 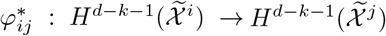 are the same. Since the persistence barcode is uniquely determined by these dimensions and the ranks (Lemma 5), we have that the corresponding persistence diagrams are equivalent 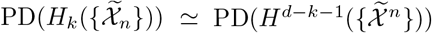. i.e.,

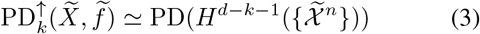

Now, to finish the proof of Step 1, we need to show equivalence of the persistent cohomology 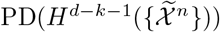 with the persistent homology for superlevel filtration 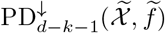. This step directly follows from the duality of persistent homology and cohomology (Lemma 4) as follows: For superlevel filtration, we use the 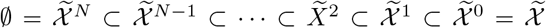. Similarly, for persistent cohomology, we use the same filtration in reverse order 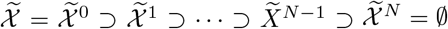. Then, by Lemma 4, their barcodes are identical:

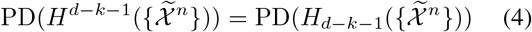

Note that to avoid crowded notation, we defined ^*n*^ with *f* (*η*) *> t*_*n*_ condition, while the original superlevel filtration has *𝒳* ^*n*^ with *f* (*η*) ≥ *t*_*n*_. Hence, in our setup *f* (*η*) *> t*_*n*_ is equivalent condition to *f* (*η*) ≥ *t*_*n*+1_. Therefore, for any 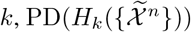 with our convention shifts each barcode by one ([*b, d*) → ([*b* +1, *d*+1)) from the original superlevel 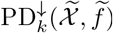. Hence, we have the equivalence:

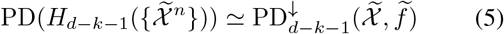

The proof of Step 1 follows from Equations 3,4,5. □

**Step 2:** 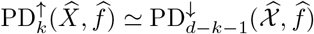

*Proof of Step 2:* After Step 1, all we need to do is to relate the homology groups in the persistence modules induced by the extended spheres 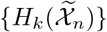 and the extended images 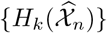.

Recall that 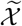 is obtained by doubling 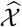. Hence, 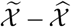 is an open ball, say *U*. By applying Mayer-Vietoris sequence to 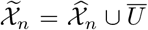, we see that 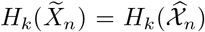 for any 0 ≤ *k d ™* 2 as 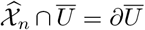 which is a (*d ™* 1)- sphere, and 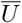 is contractible.

For *n < N*, 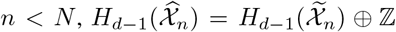 as removed ball *U* only effects the homology class corresponding to 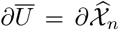 which is nontrivial in 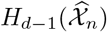 for *n < N*, while it is trivial in 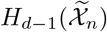 for any *n*. In Alexander duality, this homology class corresponds to the complement of the main connected component, but it is trivial in extended sphere setting. Hence, in the extended image setting, we recover this dual class in 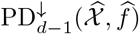 as a nontrivial homology class corresponding to the infinite bar removed sphere setting. in reduced persistence diagram 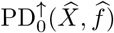 in Step 1. In other words, 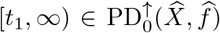 corresponds to the bar 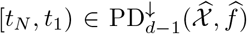 which does not exist in extended

Since other dimensions are not affected, and the correspondence in *k* = 0 is recovered as explained in the previous paragraph, the correspondence between the (unreduced) persistence diagrams in the extended image setting follows.

□

With Step 1 and Step 2, the proof of the theorem follows.

□

###### Remark 7 (Explicit Barcode Correspondence)

*In the theorem above, the explicit barcode correspondence is as follows: Let the sublevel filtration is defined as* 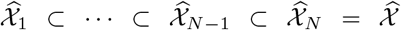 *with* 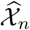 *consists of voxels* Δ_*η*_ *with f* (*η*) ≤ *t*_*n*_. *Let the superlevel filtration for the same threshold set is defined as* 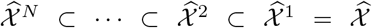 *with* 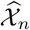 *consists of voxels* Δ_*η*_ *with f* (*η*) *t*_*n*_. *Then, if σ represents a k-dimensional topological feature in the sublevel filtration* {*χ*_*n*_} *with barcode* [*b*_*σ*_, *d*_*σ*_), *then the last time appears in the sequence at* 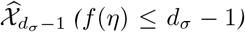. *The complement of* 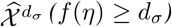 *is (f* (*η*) > *d*_*σ*_*). Hence, the corresponding (dual)* (*d – k ™* 1)*-dimensional topological feature σ*^∗^ *first time appears in* 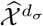 *in the sequence*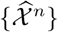, *i*.*e*., *b*_*σ* ∗_ = *d*_*σ*_. *Similarly, one can show d*_*σ*∗_ = *b*_*σ*_. *Therefore, the isomorphism* 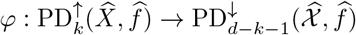 *can be defined as*

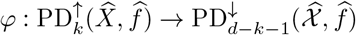

*where* φ([*b*_*σ*_, *d*_*σ*_)) = [*d*_*σ*_, *b*_*σ*_) = [*b*_*σ* ∗_, *d*_*σ*∗_) *Notice that in superlevel filtration since the nested sequence* 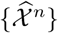 *comes with decreasing thresholds, with our convention, the birth time (b*_*σ*∗_ = *t*_*i*_*) is larger than the death time (d*_*σ*∗_ = *t*_*j*_*) in superlevel filtration. Recall that for k* = 0, *the infinite barcode* 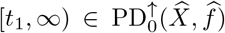 *corresponds to* 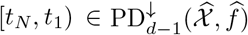 *as explained in Step 2 above*.

###### Remark 8 (Alternative extension for 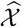)

*When defining* 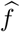 *for the extended image* 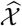, *one can use choose the value of* 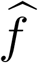 *on all boundary voxels*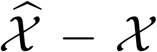 *in as* min *f instead of* max *f. The whole proof would go through by swapping sublevel and superlevel filtrations with the function ™ f instead of f. Note that the original extended image condition* 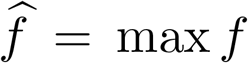 *on* 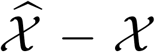 *is automatically satisfied by light background images while alternative extended image condition* 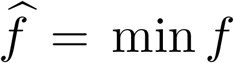 *on* 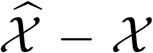 *is automatically satisfied by dark background images. This observation is important in applications of our result*.

## References

[1] Barsha Abhisheka, Saroj Kumar Biswas, and Biswajit Purkayastha. A comprehensive review on breast cancer detection, classification and segmentation using deep learning. Archives of Computational Methods in Engineering, 30(8): 5023–5052, 2023. 2

[2] Henry Adams, Tegan Emerson, Michael Kirby, Rachel Neville, Chris Peterson, Patrick Shipman, Sofya Chepushtanova, Eric Hanson, Francis Motta, and Lori Ziegelmeier. Persistence images: A stable vector representation of persistent homology. Journal of Machine Learning Research, 18 (1):218–252, 2017. 3, 13

[3] Mugahed A Al-Antari, Mohammed A Al-Masni, Mun-Taek Choi, Seung-Moo Han, and Tae-Seong Kim. A fully integrated computer-aided diagnosis system for digital x-ray mammograms via deep learning detection, segmentation, and classification. International journal of medical informatics, 117:44–54, 2018. 2

[4] Walid Al-Dhabyani, Mohammed Gomaa, Hussien Khaled, and Aly Fahmy. Dataset of breast ultrasound images. Data in brief, 28:104863, 2020. 5

[5] Dashti Ali, Aras Asaad, Maria-Jose Jimenez, Vidit Nanda, Eduardo Paluzo-Hidalgo, and Manuel Soriano-Trigueros. A survey of vectorization methods in topological data analysis. IEEE Transactions on Pattern Analysis and Machine Intelligence, 2023. 3

[6] Fouzia Altaf, Syed MS Islam, Naveed Akhtar, and Naeem Khalid Janjua. Going deep in medical image analysis: concepts, methods, challenges, and future directions. IEEE Access, 7:99540–99572, 2019. 1

[7] Gelan Ayana and Se-Woon Choe. BUVITNET: Breast ultrasound detection via vision transformers. Diagnostics, 12 (11):2654, 2022. 2

[8] Jun Bai, Russell Posner, Tianyu Wang, Clifford Yang, and Sheida Nabavi. Applying deep learning in digital breast to-mosynthesis for automatic breast cancer detection: A review. Medical image analysis, 71:102049, 2021. 2

[9] Bea Bleile, Adélie Garin, Teresa Heiss, Kelly Maggs, and Vanessa Robins. The persistent homology of dual digital image constructions. In Research in Computational Topology 2, pages 1–26. Springer, 2022. 14

[10] Magnus Botnan and Michael Lesnick. An introduction to multiparameter persistence. In Representations of Algebras and Related Structures, pages 77–150. 2023. 14

[11] Peter Bubenik and Paweł Dłotko. A persistence landscapes toolbox for topological statistics. Journal of Symbolic Computation, 78:91–114, 2017. 3

[12] Luigi Caputi, Anna Pidnebesna, and Jaroslav Hlinka. Promises and pitfalls of topological data analysis for brain connectivity analysis. NeuroImage, 238:118245, 2021. 2

[13] Mathieu Carrière, Frédéric Chazal, Yuichi Ike, Théo Lacombe, Martin Royer, and Yuhei Umeda. Perslay: A neural network layer for persistence diagrams and new graph topological signatures. In International Conference on Artificial Intelligence and Statistics, pages 2786–2796, 2020. 4

[14] Frédéric Chazal, Brittany Terese Fasy, Fabrizio Lecci, Alessandro Rinaldo, and Larry Wasserman. Stochastic convergence of persistence landscapes and silhouettes. In Proceedings of the thirtieth annual symposium on Computational geometry, pages 474–483, 2014. 3

[15] Tianqi Chen and Carlos Guestrin. Xgboost: A scalable tree boosting system. In Proceedings of the 22nd acm sigkdd international conference on knowledge discovery and data mining, pages 785–794, 2016. 4

[16] Joon Ho Choi, Hyun-Ah Kim, Wook Kim, Ilhan Lim, Inki Lee, Byung Hyun Byun, Woo Chul Noh, Min-Ki Seong, Seung-Sook Lee, Byung Il Kim, et al. Early prediction of neoadjuvant chemotherapy response for advanced breast cancer using pet/mri image deep learning. Scientific reports, 10(1):21149, 2020. 2

[17] James R Clough, Nicholas Byrne, Ilkay Oksuz, Veronika A Zimmer, Julia A Schnabel, and Andrew P King. A topological loss function for deep-learning based image segmentation using persistent homology. IEEE transactions on pattern analysis and machine intelligence, 44(12):8766–8778, 2020. 2

[18] Baris Coskunuzer and Cüneyt Gürcan Akçora. Topological methods in machine learning: A tutorial for practitioners. arXiv preprint 2409.02901, 2024. 2

[19] Lorin Crawford, Anthea Monod, Andrew X Chen, Sayan Mukherjee, and Raúl Rabadán. Predicting clinical outcomes in glioblastoma: an application of topological and functional data analysis. Journal of the American Statistical Association, 115(531):1139–1150, 2020. 2

[20] Qing Dan, Ziting Xu, Hannah Burrows, Jennifer Bissram, Jeffrey SA Stringer, and Yingjia Li. Diagnostic performance of deep learning in ultrasound diagnosis of breast cancer: a systematic review. NPJ Precision Oncology, 8(1):21, 2024. 2

[21] Vin De Silva, Dmitriy Morozov, and Mikael Vejdemo-Johansson. Dualities in persistent (co) homology. Inverse Problems, 27(12):124003, 2011. 14

[22] Walter H Delashmit, Michael T Manry, et al. Recent developments in multilayer perceptron neural networks. In Proceedings of the seventh annual memphis area engineering and science conference, MAESC, page 33, 2005. 4

[23] Karin Dembrower, Erik Wåhlin, Yue Liu, Mattie Salim, Kevin Smith, Peter Lindholm, Martin Eklund, and Fredrik Strand. Effect of artificial intelligence-based triaging of breast cancer screening mammograms on cancer detection and radiologist workload: a retrospective simulation study. The Lancet Digital Health, 2(9):e468–e474, 2020. 2

[24] Tamal Krishna Dey and Yusu Wang. Computational topology for data analysis. Cambridge University Press, 2022. 2

[25] Tomoyuki Fujioka, Mio Mori, Kazunori Kubota, Jun Oyama, Emi Yamaga, Yuka Yashima, Leona Katsuta, Kyoko Nomura, Miyako Nara, Goshi Oda, et al. The utility of deep learning in breast ultrasonic imaging: a review. Diagnostics, 10(12):1055, 2020. 2

[26] Behnaz Gheflati and Hassan Rivaz. Vision transformers for classification of breast ultrasound images. In IEEE EMBC, pages 480–483. IEEE, 2022. 2

[27] Jean Dickinson Gibbons and Subhabrata Chakraborti. Nonparametric statistical inference. CRC press, 2014. 8

[28] Barbara Giunti. TDA applications library, 2022. https://www.zotero.org/groups/2425412/tda-applications/library. 2

[29] Barbara Giunti, Janis Lazovskis, and Bastian Rieck. DONUT: Database of Original & Non-Theoretical Uses of Topology, 2022. https://donut.topology.rocks. 2

[30] Wilfrido Gómez-Flores, Maria Julia Gregorio-Calas, and Wagner Coelho de Albuquerque Pereira. Bus-bra: A breast ultrasound dataset for assessing computer-aided diagnosis systems. Medical Physics, 2023. 5

[31] Mustafa Hajij, Ghada Zamzmi, and Fawwaz Batayneh. Tdanet: fusion of persistent homology and deep learning features for covid-19 detection from chest x-ray images. In 2021 43rd Annual International Conference of the IEEE Engineering in Medicine & Biology Society (EMBC), pages 4115–4119. IEEE, 2021. 2

[32] Ghada Hamed, Mohammed Abd El-Rahman Marey, Safaa El-Sayed Amin, and Mohamed Fahmy Tolba. Deep learning in breast cancer detection and classification. In Proceedings of the International Conference on Artificial Intelligence and Computer Vision (AICV2020), pages 322–333. Springer, 2020. 2

[33] Allen Hatcher. Algebraic Topology. Cambridge University Press, 2002. 14, 15

[34] Kaiming He et al. Deep residual learning for image recognition. In CVPR, pages 770–778, 2016. 5

[35] Felix Hensel, Michael Moor, and Bastian Rieck. A survey of topological machine learning methods. Frontiers in Artificial Intelligence, 4:52, 2021. 1

[36] Essam H Houssein, Marwa M Emam, Abdelmgeid A Ali, and Ponnuthurai Nagaratnam Suganthan. Deep and machine learning techniques for medical imaging-based breast cancer: A comprehensive review. Expert Systems with Applications, 167:114161, 2021. 2

[37] Andrew G Howard, Menglong Zhu, Bo Chen, Dmitry Kalenichenko, Weijun Wang, Tobias Weyand, Marco Andreetto, and Hartwig Adam. Mobilenets: Efficient convolutional neural networks for mobile vision applications. arXiv preprint 1704.04861, 2017. 5

[38] Gao Huang, Zhuang Liu, Laurens Van Der Maaten, and Kilian Q Weinberger. Densely connected convolutional networks. In CVPR, pages 4700–4708, 2017. 5

[39] Johanna Immonen, Amauri Souza, and Vikas Garg. Going beyond persistent homology using persistent homology. Advances in Neural Information Processing Systems, 36, 2024. 1

[40] Sohail Iqbal, H Fareed Ahmed, Talha Qaiser, Muhammad Imran Qureshi, and Nasir Rajpoot. Classification of covid-19 via homology of ct-scan. arXiv preprint 2102.10593, 2021. 2

[41] Lida Kanari, Paweł Dłotko, Martina Scolamiero, Ran Levi, Julian Shillcock, Kathryn Hess, and Henry Markram. A topological representation of branching neuronal morphologies. Neuroinformatics, 16(1):3–13, 2018. 2

[42] Ekaterina Khramtsova, Guido Zuccon, Xi Wang, and Mahsa Baktashmotlagh. Rethinking persistent homology for visual recognition. In Topological, Algebraic and Geometric Learning Workshops 2022, pages 206–215. PMLR, 2022. 2

[43] Jieun Koh, Youngno Yoon, Sungwon Kim, Kyunghwa Han, and Eun-Kyung Kim. Deep learning for the detection of breast cancers on chest computed tomography. Clinical breast cancer, 22(1):26–31, 2022. 2

[44] Geert Litjens, Thijs Kooi, Babak Ehteshami Bejnordi, Arnaud Arindra Adiyoso Setio, Francesco Ciompi, Mohsen Ghafoorian, Jeroen Awm Van Der Laak, Bram Van Ginneken, and Clara I Sánchez. A survey on deep learning in medical image analysis. Medical Image Analysis, 42:60–88, 2017. 1

[45] Ze Liu, Yutong Lin, Yue Cao, Han Hu, Yixuan Wei, Zheng Zhang, Stephen Lin, and Baining Guo. Swin transformer: Hierarchical vision transformer using shifted windows. In Proceedings of the IEEE/CVF international conference on computer vision, pages 10012–10022, 2021. 4

[46] Ze Liu, Han Hu, Yutong Lin, Zhuliang Yao, Zhenda Xie, Yixuan Wei, Jia Ning, Yue Cao, Zheng Zhang, Li Dong, et al. Swin transformer v2: Scaling up capacity and resolution. In Proceedings of the IEEE/CVF conference on computer vision and pattern recognition, pages 12009–12019, 2022. 4, 5, 6

[47] Melissa R McGuirl, Alexandria Volkening, and Björn Sandstede. Topological data analysis of zebrafish patterns. Proceedings of the National Academy of Sciences, 117(10): 5113–5124, 2020. 2

[48] Maged Nasser and Umi Kalsom Yusof. Deep learning based methods for breast cancer diagnosis: A systematic review and future direction. Diagnostics, 13(1):161, 2023. 2

[49] Theodore Papamarkou, Tolga Birdal, Michael Bronstein, Gunnar Carlsson, Justin Curry, Yue Gao, Mustafa Hajij, Roland Kwitt, Pietro Liò, Paolo Di Lorenzo, et al. Position paper: Challenges and opportunities in topological deep learning. arXiv preprint 2402.08871, 2024. 2

[50] Yaopeng Peng, Hongxiao Wang, Milan Sonka, and Danny Z Chen. Phg-net: Persistent homology guided medical image classification. In Proceedings of the IEEE/CVF Winter Conference on Applications of Computer Vision, pages 7583–7592, 2024. 2, 12

[51] Talha Qaiser, Yee-Wah Tsang, Daiki Taniyama, Naoya Sakamoto, Kazuaki Nakane, David Epstein, and Nasir Rajpoot. Fast and accurate tumor segmentation of histology images using persistent homology and deep convolutional features. Medical image analysis, 55:1–14, 2019. 2

[52] Xuejun Qian, Jing Pei, Hui Zheng, Xinxin Xie, Lin Yan, Hao Zhang, Chunguang Han, Xiang Gao, Hanqi Zhang, Weiwei Zheng, et al. Prospective assessment of breast cancer risk from multimodal multiview ultrasound images via clinically applicable deep learning. Nature biomedical engineering, 5 (6):522–532, 2021. 2

[53] Xiaolei Qu, Hongyan Lu, Wenzhong Tang, Shuai Wang, Dezhi Zheng, Yaxin Hou, and Jue Jiang. A vgg attention vision transformer network for benign and malignant classification of breast ultrasound images. Medical Physics, 49(9): 5787–5798, 2022. 2

[54] Raúl Rabadán and Andrew J Blumberg. Topological data analysis for genomics and evolution: topology in biology. Cambridge University Press, 2019. 2

[55] Bastian Rieck, Tristan Yates, Christian Bock, Karsten Borgwardt, Guy Wolf, Nicholas Turk-Browne, and Smita Krishnaswamy. Uncovering the topology of time-varying fMRI data using cubical persistence. NeurIPS, 33:6900–6912, 2020. 2

[56] Paulo Sergio Rodrigues. Breast ultrasound image. Mendeley Data, 1(10.17632), 2017. 5

[57] Roslidar Roslidar, Aulia Rahman, Rusdha Muharar, Muhammad Rizky Syahputra, Fitri Arnia, Maimun Syukri, Biswajeet Pradhan, and Khairul Munadi. A review on recent progress in thermal imaging and deep learning approaches for breast cancer detection. IEEE Access, 8:116176–116194, 2020. 2

[58] Ainkaran Santhirasekaram, Mathias Winkler, Andrea Rockall, and Ben Glocker. Topology preserving compositionality for robust medical image segmentation. In CVPR, pages 543–552, 2023. 2

[59] Shallu Sharma and Rajesh Mehra. Conventional machine learning and deep learning approach for multi-classification of breast cancer histopathology images—a comparative insight. Journal of digital imaging, 33:632–654, 2020. 2

[60] Dinggang Shen, Guorong Wu, and Heung-Il Suk. Deep learning in medical image analysis. Annual review of biomedical engineering, 19:221–248, 2017. 1

[61] Li Shen, Laurie R Margolies, Joseph H Rothstein, Eugene Fluder, Russell McBride, and Weiva Sieh. Deep learning to improve breast cancer detection on screening mammography. Scientific reports, 9(1):12495, 2019. 2

[62] Yiqiu Shen, Farah E Shamout, Jamie R Oliver, Jan Witowski, Kawshik Kannan, Jungkyu Park, Nan Wu, Connor Huddleston, Stacey Wolfson, Alexandra Millet, et al. Artificial intelligence system reduces false-positive findings in the interpretation of breast ultrasound exams. Nature communications, 12(1):5645, 2021. 2

[63] K Simonyan and A Zisserman. Very deep convolutional networks for large-scale image recognition. In ICLR, 2015. 5

[64] Amitojdeep Singh, Sourya Sengupta, and Vasudevan Lakshminarayanan. Explainable deep learning models in medical image analysis. Journal of imaging, 6(6):52, 2020. 1

[65] Yashbir Singh et al. TDA in Medical Imaging: Current state of the art. Insights into Imaging, 14(1):1–10, 2023. 1, 2

[66] Ann E Sizemore, Jennifer E Phillips-Cremins, Robert Ghrist, and Danielle S Bassett. The importance of the whole: topological data analysis for the network neuroscientist. Network Neuroscience, 3(3):656–673, 2019. 1

[67] Yara Skaf and Reinhard Laubenbacher. Topological data analysis in biomedicine: A review. Journal of Biomedical Informatics, page 104082, 2022. 2

[68] Eashwar Somasundaram, Adam Litzler, Raoul Wadhwa, Steph Owen, and Jacob Scott. Persistent homology of tumor ct scans is associated with survival in lung cancer. Medical physics, 48(11):7043–7051, 2021. 2

[69] N. Stucki et al. Topologically faithful image segmentation via induced matching of persistence barcodes. In ICML, 2023. 2

[70] Hyuna Sung, Jacques Ferlay, Rebecca L Siegel, Mathieu Laversanne, Isabelle Soerjomataram, Ahmedin Jemal, and Freddie Bray. Global cancer statistics 2020: Globocan estimates of incidence and mortality worldwide for 36 cancers in 185 countries. CA: a cancer journal for clinicians, 71(3): 209–249, 2021. 1

[71] Christian Szegedy, Sergey Ioffe, Vincent Vanhoucke, and Alexander Alemi. Inception-v4, inception-resnet and the impact of residual connections on learning. In Proceedings of the AAAI conference on artificial intelligence, 2017. 5

[72] Mingxing Tan and Quoc Le. Efficientnet: Rethinking model scaling for convolutional neural networks. In ICML, pages 6105–6114. PMLR, 2019. 5

[73] A Vaswani et al. Attention is all you need. Advances in Neural Information Processing Systems, 2017. 4

[74] Fan Wang, Saarthak Kapse, Steven Liu, Prateek Prasanna, and Chao Chen. Topotxr: a topological biomarker for predicting treatment response in breast cancer. In International Conference on Information Processing in Medical Imaging, pages 386–397. Springer, 2021. 2

[75] Chi-Chong Wong and Chi-Man Vong. Persistent homology based graph convolution network for fine-grained 3D shape segmentation. In ICCV, pages 7098–7107, 2021. 2

[76] Ankur Yadav, Faisal Ahmed, Ovidiu Daescu, Reyhan Gedik, and Baris Coskunuzer. Histopathological cancer detection with topological signatures. In 2023 IEEE International Conference on Bioinformatics and Biomedicine (BIBM), pages 1610–1619. IEEE, 2023. 2

[77] Zuoyu Yan and Tengfei Ma. Neural approximation of extended persistent homology on graphs. In Advances in Neural Information Processing Systems, 2022. 1

[78] Jiancheng Yang, Rui Shi, Donglai Wei, Zequan Liu, Lin Zhao, Bilian Ke, Hanspeter Pfister, and Bingbing Ni. Medmnist v2-a large-scale lightweight benchmark for 2D and 3D biomedical image classification. Scientific Data, 10(1):41, 2023. https://medmnist.com. 5, 7, 12

[79] Xujing Yao, Xinyue Wang, Shui-Hua Wang, and Yu-Dong Zhang. A comprehensive survey on convolutional neural network in medical image analysis. Multimedia Tools and Applications, 81(29):41361–41405, 2022. 1

[80] Ali Zia, Abdelwahed Khamis, James Nichols, Zeeshan Hayder, Vivien Rolland, and Lars Petersson. Topological deep learning: A review of an emerging paradigm. arXiv preprint 2302.03836, 2023. 2

